# Genetic determinants of Multiple Sclerosis susceptibility in diverse ancestral backgrounds

**DOI:** 10.1101/2025.01.16.25320672

**Authors:** Benjamin M. Jacobs, Luisa Schalk, Emily Tregaskis-Daniels, Antonio Scalfari, Ashwini Nandoskar, Angie Dunne, Bruno Gran, Charles A. Mein, Charlotte Sellers, Cord Spilker, David Rog, Elisa Visentin, Elizabeth Lindsey Bezzina, Emeka Uzochukwu, Emma Tallantyre, Eva Wozniak, Eve Sacre, Ghaniah Hassan-Smith, Helen L. Ford, Jade Harris, Joan Bradley, Joshua Breedon, Judith Brooke, Karim L. Kreft, Katila George, Maria Papachatzaki, Martin O’Malley, Michelle Peter, Miriam Mattoscio, Neisha Rhule, Nikos Evangelou, Nimisha Vinod, Outi Quinn, Ramya Shamji, Rashmi Kaimal, Rebecca Boulton, Riffat Tanveer, Rod Middleton, Roxanne Murray, Ruth Bellfield, Sadid Hoque, Shakeelah Patel, Sonia Raj, Stephanie Gumus, Stephanie Mitchell, Stephen Sawcer, Tarunya Arun, Tatiana Pogreban, Terri-Louise Brown, Thamanna Begum, Veronica Antoine, Waqar Rashid, Grace Fawehinmi, Claire Reidy, Shanaz Begum, Shannon Bernard Healey, Harriet Cummins, Kelly Westwood, Deborah Spencer, Shegufta Farooq, Katharine Harding, Sarah Williams, Georgina Radford, Selina White, Nathan Alldred-Douglas, Linford Fernandes, Adil Harroud, Jacob L. McCauley, Ashley Beecham, Nicolas Vince, Nayane dos Santos Brito Silva, Huw R Morris, Eli Silber, Gavin Giovannoni, Alastair J. Noyce, Ruth Dobson

## Abstract

The genetic architecture of Multiple Sclerosis (MS) susceptibility has been extensively assessed in populations of European ancestry. Greater ancestral diversity in genetic analyses of MS susceptibility is needed to improve the utility of Multiple Sclerosis genetic risk scores, fine map causal variants underlying established associations, and thereby enhance the identification of drug targets. Here we report findings from a genetic study of Multiple Sclerosis susceptibility in an ancestrally-diverse United Kingdom-based cohort.

Participants with Multiple Sclerosis were recruited via clinical sites, an online platform, and through the United Kingdom Multiple Sclerosis Register. Phenotype data were gathered using a standardised questionnaire. DNA was extracted from saliva samples obtained remotely or in person, and participants were genotyped using a commercial genotyping array. Following imputation, cases were combined with controls from the United Kingdom Biobank and subjected to stringent quality control and genetic ancestry inference. We defined two broad ancestral groups of South Asian and African ancestry. We performed within-ancestry case-control genome-wide association studies of Multiple Sclerosis susceptibility using logistic models accounting for population structure and sex. We examined both single nucleotide variants and imputed classical Human Leukocyte Antigen alleles.

We curated two ancestrally-matched case-control genetic datasets (South Asian ancestry: N_Case_=175, N_Control_=6744; African ancestry: N_Case_=113, N_Control_=5177). In both ancestries, we found genetic variants within the Major Histocompatibility Complex associated with Multiple Sclerosis susceptibility (South Asian ancestry: lead variant chr6:32600515:G:A on hg38 co-ordinates, Odds Ratio=1.84, nearest gene *HLA-DRB1*, *P=*4.6×10^−6^; African ancestry: lead variant chr6:29919337:A:G, Odds Ratio=2.24, nearest gene *HLA-A P=*4.3×10^−5^). European-ancestry susceptibility alleles were over-represented in cases from both ancestries, with the degree of concordance stronger for the South Asian (ρ=0.31, *P*=8.1×10^−6^) than African (ρ=0.1, *P*=0.3) ancestry cohort. European-derived genetic risk scores performed better than chance but less well than in European ancestry cohorts, explaining 1.6% (South Asian*, P*=1.0×10^−4^) and 0.5% (African*, P*=0.08) of the liability to MS.

The genetic architecture of MS susceptibility shows strong concordance across ancestral groups suggesting shared disease mechanisms. Larger studies in diverse populations are likely to enhance our understanding of how genetic variation contributes to MS susceptibility in people of all ancestral backgrounds.

## Introduction

The heritable component of Multiple Sclerosis (MS) susceptibility was recognised long before the era of modern genomics. Over the past 20 years, case-control Genome-Wide Association Studies (GWAS) orchestrated by the International Multiple Sclerosis Genetics Consortium (IMSGC) have shown that this heritable component is highly polygenic, underscored by moderately large effects of allelic variants in the Major Histocompatibility Complex (MHC), and small effects of common and low-frequency variants outside the MHC^1–6^. Most recently, case-control GWAS of a total of 47,429 MS cases and 68,374 controls expanded the number of disease-associated loci reaching genome wide statistical significance to 233, including 201 outside the MHC^1^.

International GWAS efforts have focused on populations of European ancestry. GWAS of MS susceptibility in non-European ancestry populations have been substantially more limited in sample size, and therefore statistical power^7–13^. These studies – reviewed elsewhere^14^– have broadly shown concordance in the genetic architecture of MS between ancestries, with European risk alleles tending to be over-represented in cases from all tested ancestral groups^10,12,15,16^. Studying the genetic underpinnings of MS susceptibility in diverse ancestral populations is essential to ensure that genetic risk scores for predicting MS perform equitably across populations^15–18^. This approach is also needed to refine our understanding of how genetic variation contributes to disease susceptibility through multi-ancestry fine mapping: this has proven powerful across complex traits and diseases^19–22^. In MS, genetic analysis in diverse ancestries has produced some notable successes, such as the disentangling of the *HLA-DRB1*15:01*-*DQB1*06:02* haplotype in African Americans^23–25^ and the discovery of a mechanistic link between a genetic variant and autoimmunity risk via soluble B-cell activating factor (BAFF) expression in Sardinia^26^.

In this paper we report the characteristics of an ancestrally-diverse phenotyped and genotyped UK MS cohort. We describe a multi-ancestry genetic study of MS susceptibility performed by combining this cohort with ancestrally-similar controls from UK Biobank. Our aims were to (i) explore the genetic basis of MS susceptibility in UK MS cases of South Asian and African ancestry, and (ii) quantify the degree of overlap between the genetic architecture of MS in European and non-European ancestral backgrounds, both within and outside of the Major Histocompatibility Complex locus.

## Materials and methods

An overview of the study design is given in **figure 1**.

**Figure 1:**
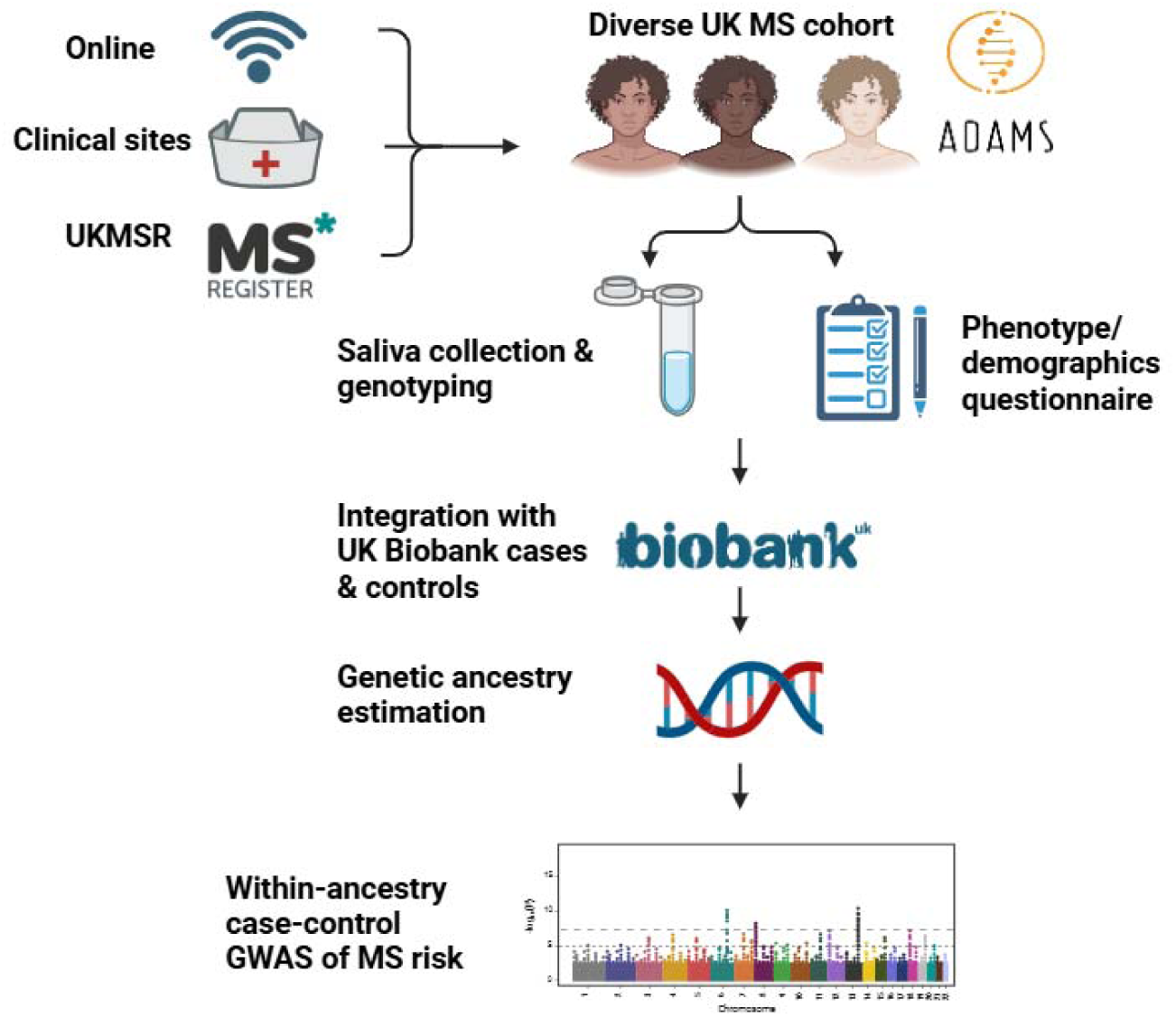
an overview of the study design. We assembled a cohort of ancestrally-diverse people with MS living in the UK. Cases were genotyped from saliva, and phenotype via a structured questionnaire. Genetic data from our case-only cohort were combined with genotyping data from the United Kingdom Biobank (UKB), which served as a source of a large number of ancestrally-diverse controls and a small number of ancestrally-diverse additional cases. Following genetic ancestry estimation, we performed within-ancestry genetic analysis of MS susceptibility.

### Cohort design & recruitment

The ADAMS project (A Genetic <u>A</u>ssociation study in <u>D</u>iverse <u>A</u>ncestries of <u>M</u>ultiple <u>S</u>clerosis) is a genotype-phenotype cohort study of individuals with MS from diverse ancestral backgrounds living in the UK^27^ (https://app.mantal.co.uk/adams, REC reference 21/PR/1289). Participants with self-reported or clinically diagnosed Multiple Sclerosis were recruited via a bespoke online platform, clinical sites, primary care, and the UK MS Register (UKMSR)^28–31^. A list of participating sites is given in **supplementary table 1**.

### Phenotype data

We collected baseline phenotypic data via a questionnaire (administered online, in person, or approximated from the UKMSR questionnaire). This captured data on participant demographics (age, gender, self-reported ethnicity), MS history (age at diagnosis, MS subtype, current and previous disease-modifying therapy), and MS risk factors (family history, country of birth, migration age if born outside the UK, Body Mass Index [BMI] during adolescence, smoking history, and glandular fever). In addition, we collected data to estimate physical disability: Participants recruited via the ADAMS website were asked to complete an online version of the Expanded Disability Status Scale (EDSS)^32^, the Multiple Sclerosis Impact Scale-29 (MSIS-29)^33^, and the EQ5D health-related quality of life scale^34^. For participants recruited via the UKMSR, we collected the most recent MSIS-29 and EDSS scores. For participants recruited in person via clinical sites, we collected EDSS scores only. EDSS scores were transformed to the global age-related Multiple Sclerosis Severity Score (gARMSS) score to account for confounding by age at ascertainment^35^.

### Genetic data, identifying controls, and ancestry inference

A detailed overview of genetic data quality control (QC) is provided in the supplementary data (**supplementary figures 1 – 11**). Briefly, we obtained genomic DNA from our cohort using saliva sampling with the Oragene OG-600 saliva kits. We performed genotyping using the Illumina Global Screening Array version 3.0. Following initial quality control of individuals and variants, we imputed our dataset to the Haplotype Reference Consortium (HRC version 1.1) using the Michigan Imputation Server – although dominated by individuals of European ancestry, the HRC panel incorporates individuals of African (N = 661) and South Asian (N = 489) ancestry from the 1,000 Genomes project^36,37^. Following further QC of the imputed data, we combined our case-only cohort with imputed genetic data from the United Kingdom Biobank (UKB^38^, application ID 78867). We inferred the genetic ancestry of cases and controls using a random forest classifier trained on reference data from the Human Genome Diversity Project^39^ and 1,000 Genomes (HGDP-1KG)^40^. Using these PCA-based inferred ancestry calls, we divided the cohort into people of inferred South Asian (SAS) and African (AFR) genetic ancestry (see **supplementary figures 3-9** and supplementary methods section ‘Ancestry Inference’ for further details). We then performed further within-ancestry quality (see supplementary methods section ‘within-ancestry quality control’ and **supplementary figure 11**) control to identify and remove batch effects and re-imputed the joint case-control SAS and AFR cohorts - comprising our cohort, UKB cases and UKB controls - using the 1,000 Genomes reference panel.

### Genome-wide association study of MS susceptibility

We conducted within-ancestry GWAS of MS susceptibility using mixed logistic models implemented in REGENIE^41^. As the ADAMS cohort is substantially younger than UKB (**supplementary figure 12**), age was highly collinear with case-control status, so we did not adjust for age in the primary analyses. We adjusted for sex and the first ten genetic principal components. Continuous covariates were standardised to have a mean of 0 and a standard deviation 1. For step 1 of REGENIE we used a lightly LD-pruned set of genome-wide high-quality markers (Imputation R^2^ > 0.97, pruning R^2^ 0.9, MAF > 5%, missingness < 1%, HWE P > 10^−10^). We tested variants down to a MAF of 5% in each case-control subset. We estimated genomic inflation using the genome-wide lambda statistic (λ = median observed χ^2^ / median expected χ^2^ from the null distribution). We included age as a covariate in sensitivity analyses and used fixed-effects logistic regression models (implemented in PLINK^42^) adjusted for age, sex, and the first ten PCs (**supplementary figures 13 – 15**).

### Variant annotation

We identified independent SNPs by LD-clumping within each ancestral group using a clumping R^2^ of 0.01 (i.e. considering all signals in LD above this threshold as a single locus; the SNP with the lowest *P* value considered to be the lead SNP). GWAS results were annotated using the Variant Effect Predictor (VEP) to determine the nearest gene and the impact of the variant on gene and transcript function^43^. All annotation was performed in hg38 co-ordinates using the C:P:R:A (chromosome: position: reference: alternate) format for variant identification. Allele frequencies were inferred from gnomAD^44^.

### Comparison with European-ancestry GWAS

We contrasted the South Asian and African-ancestry GWAS with European-ancestry GWAS using the published IMSGC discovery-phase GWAS summary statistics^1^. Summary statistics were merged based on strict matching of C:P:R:A coordinates. We first filtered these variants to those present in the SAS- and AFR-ancestry GWAS. To determine the concordance of effect sizes, we defined independent SNPs in the European-ancestry GWAS using LD-clumping, defining independent SNPs by searching within 1MB windows around all SNPs with *P* < 5×10^−8^, and combining those with a pairwise R^2^ > 0.01. Clumping was performed using the 503 European ancestry individuals from the 1000 Genomes project. Concordance was defined using a one-tailed Binomial test considering whether the established European-ancestry variant SNP was risk-increasing or protective in the SAS/AFR GWAS. Under the null hypothesis the probability of observing the same effect direction as the European-ancestry GWAS is 50% for each SNP.

### HLA allelic association

We used two approaches to impute and fine-map the association signals at the Major Histocompatibility Complex (MHC) locus on chromosome 6. For the primary analysis we imputed classical HLA alleles at six loci (*HLA-A, B, C, DQB1, DRB1,* and *DPB1*) using the HIBAG R package^45,46^. We used classifiers trained on a multi-ancestry reference panel (provided by the SNP-HLA Reference Consortium [SHLARC] consortium)^47,48^. We imputed HLA alleles to 4-digit (two-field) resolution, i.e. to the level of nonsynonymous variants affecting the sequence. We then validated these HLA allele calls using a phasing-based approach, SNP2HLA^49^ (with the 1,000 Genomes dataset as reference). We tested for association between common (MAF >= 1%) classical HLA alleles and MS risk within each ancestry using logistic regression models adjusted for the same covariates as GWAS (sex, PCs 1-10). We performed testing at 4-digit resolution using an additive genetic model (i.e. encoding the number of copies of each allele as 0/1/2). Global HLA allele frequencies were obtained via the Allele Frequency Net Database^50^. In addition, HLA allele frequencies were contrasted with those derived via an alternative method (HLA*IMP) by UK Biobank (data field 22182)^51^ in the entire genotyped UK Biobank dataset. We calculated LD between classical HLA alleles and defined haplotypes using the phased output from SNP2HLA to compute D’ and R^2^, and the frequency of overall long-range haplotypes spanning all six genes assessed. We performed stepwise conditional analysis to determine independent HLA allelic associations in R (using the SNP2HLA imputation). HLA associations were contrasted with previous studies in European ancestries^1,6,52^, South Asian ancestry^11^, and African Americans^9^.

The population attributable fraction (PAF) of HLA-DRB1*15:01 was estimated using both Levin’s and Miettinen’s approximations^53,54^, i.e.:

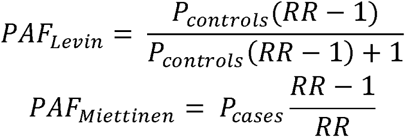

In both cases *P* indicates the prevalence of the risk factor (i.e. HLA-DRB1*15:01), and *RR* indicates the Relative Risk, approximated by the Odds Ratio. We have also made the simplifying assumption of a dominant model for these calculations (i.e. OR_Dominant_ = OR_Additive_), and approximated the prevalence of carriage using Hardy-Weinberg equilibrium (i.e. prevalence of DRB1*15:01 = *p*^2^ + 2*pq*, where *p* = allele frequency).

### Polygenic risk score profiling

We derived risk scores using common (MAF > 5%) high-quality variants from the IMSGC discovery-phase GWAS meta-analysis of 14,802 cases and 26,703 controls of European ancestry^1^. PRS calculation and model fitting was performed using the clumping-and-thresholding approach implemented in PRSice-2^55^. Briefly, this approach calculated a weighted score for each individual based on the number of risk alleles they carry, with the contribution of each allele weighted by the effect size in the reference (European-ancestry) GWAS. To define a set of high-quality markers we used the intersection of IMSGC GWAS SNPs and the SNPs we used for GWAS. We defined several scores by varying the *P* value threshold for SNP inclusion and the clumping R^2^ value used to define independent loci. We used in-sample LD within each ancestry to define independent variants. We evaluated the performance of each PRS using logistic regression models adjusted for the same covariates as in the GWAS (sex, PCs 1-10).

### Ethical approval

The London—South East Research Ethics Committee (REC reference 21/PR/1289) has approved this study and its amendments.

### Computing

Analysis was conducted using the Apocrita High Performance Computing (HPC) cluster provided by Queen Mary University of London^56^.

## Results

### Cohort description

We recruited and genotyped a cohort of people with Multiple Sclerosis (MS) living in the UK with an emphasis on people from diverse ancestral backgrounds (**figure 1**). We restricted the dataset to those who had returned a saliva kit, provided high-quality DNA, and passed genetic quality control (final N = 736, **supplementary figure 1**). The demographic and phenotypic characteristics of the cohort were typical of other modern MS cohorts in terms of age at diagnosis, gender balance, and disease characteristics (**table 1**). The genotyped cohort (N = 736) comprised people of self-reported South Asian (N = 186), Black (N = 122), Mixed/Other (N = 143), and White (N = 223) ethnic backgrounds, as well as people with missing ethnicity data (N = 62).

**Table 1:**
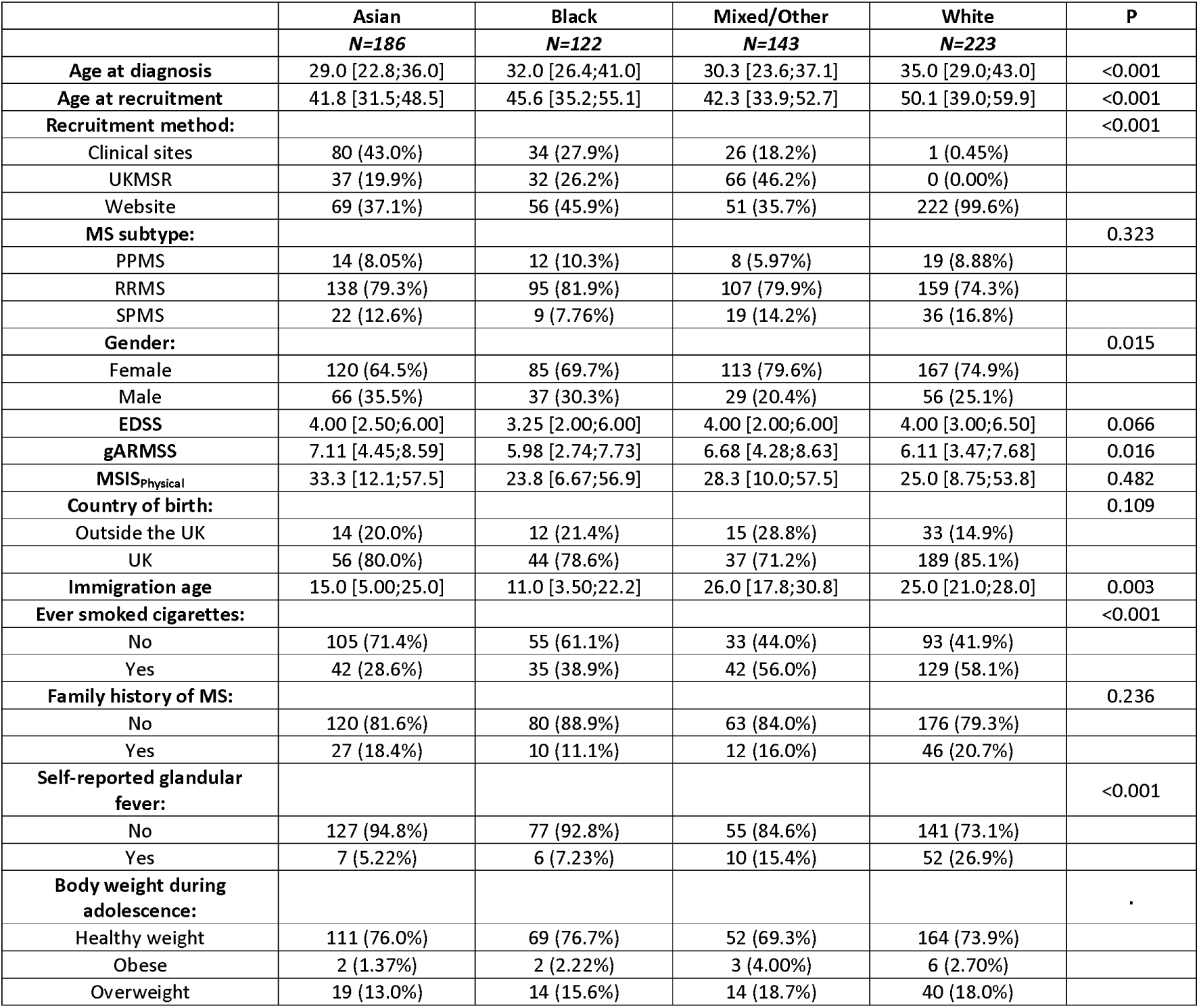
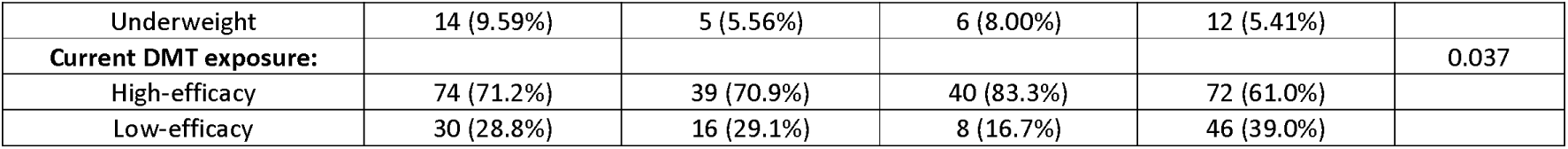
demographic characteristics of the genotyped cohort, split by self-reported ethnic background. Continuous variables are presented as the median and interquartile range; categorical variables are presented as n and %. Missing data within each category were excluded, i.e. the denominator for each variable is those with complete data.

### Genetic ancestry inference

We combined genetic data from our case-only cohort with UK Biobank, estimated the genetic ancestry of cases and controls in this combined cohort, and identified groups of ancestrally similar cases and controls (**figures 2A & B; supplementary figures 3 - 9**). After the exclusion of individuals with ambiguous genetic ancestry, we focussed our analysis on people of inferred African (AFR) ancestry (112 ADAMS cases, 17 UKB cases, 7701 UKB controls) and people of South Asian (SAS) ancestry (201 ADAMS cases, 13 UKB cases, 9001 UKB controls). These genetic ancestry estimates corresponded broadly with self-reported ethnicity (**figures 2C & 2D**).

**Figure 2:**
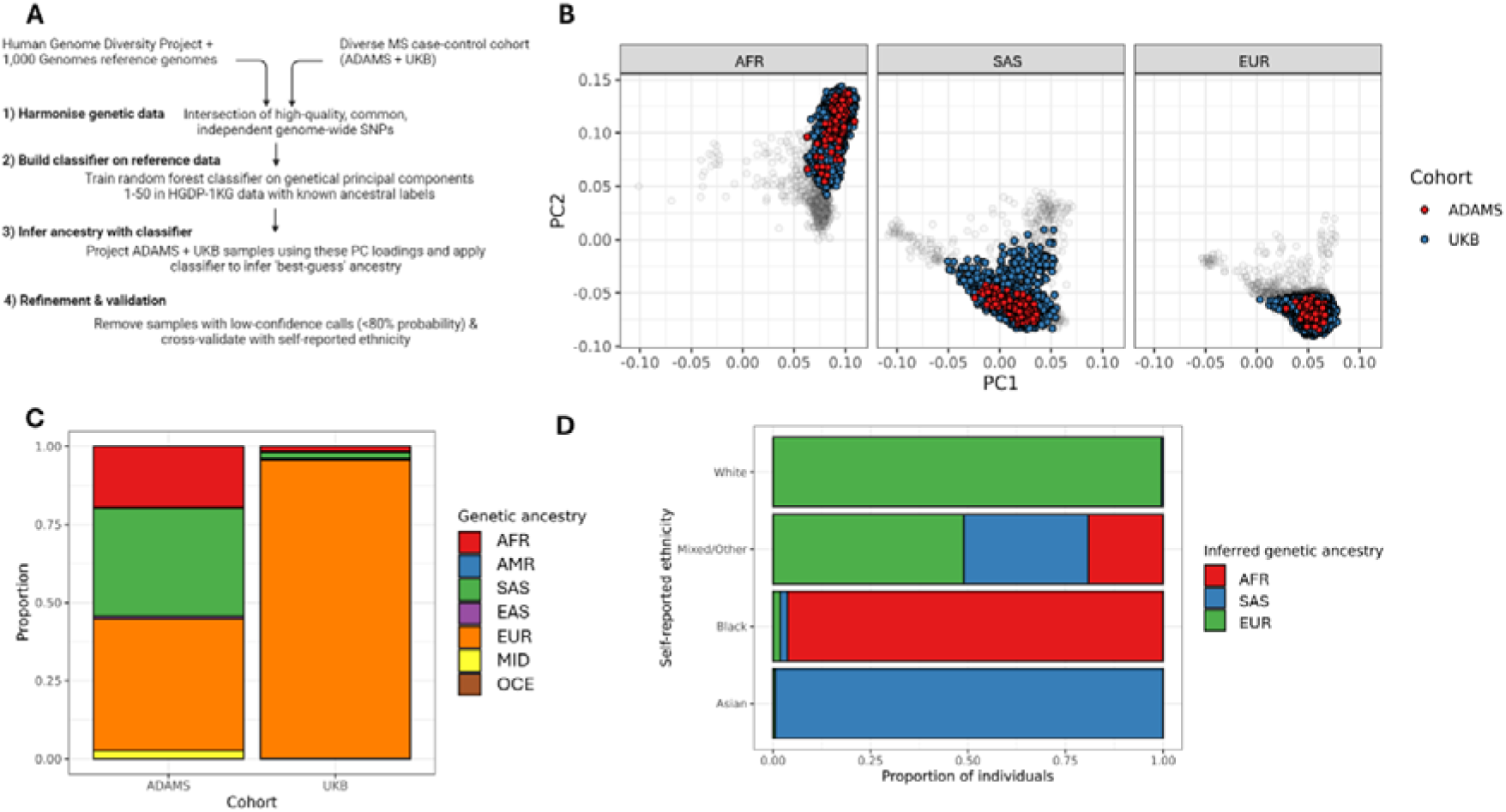
Genetic ancestry estimation and identification of ancestrally-similar cases & controls. A: an overview of the strategy used to infer genetic ancestry. B: PCA plots showing the first two genetic principal components of the joint UKB-ADAMS dataset. Panels show (in order) participants of inferred African (AFR), South Asian (SAS), and European (EUR) ancestry, coloured by cohort. Greyed out dots indicate participants assigned to the ancestry group with low confidence (probability <80%), who were excluded from downstream analysis. C: Stacked barplots showing the proportion of each cohort of each broad continental genetic ancestry group. D: horizontal stacked barplots showing the correspondence between self-reported ethnicity and genetic ancestry in the ADAMS cohort.

### MS Susceptibility GWAS in South Asian and African ancestries

Following further quality control, including removal of ancestral outliers, we performed within-ancestry case-control genome-wide association studies (GWAS) of MS susceptibility in people of genetically-inferred South Asian (SAS; N_MS_ = 175, N_Control_ = 6744) and African (AFR; N_MS_ = 113, N_Control_ = 5177) ancestry. In both SAS and AFR ancestries there were nominally significant signals (*P* < 1×10^−4^) at the Major Histocompatibility Complex (MHC) locus (chromosome 6, BP 25,000,000 – 35,000,000), consistent with this locus harbouring the strongest genetic susceptibility signals for MS across populations (**figure 3**, **table 2**). Outside the MHC region, we identified a total of three (SAS) and eighteen (AFR) independent loci showing nominally significant association with MS risk (*P* < 1×10^−4^). Other than the MHC signals, none of the other suggestive loci were within 1MB of a suggestive European-ancestry MS risk allele (**figure 3**, **table 2**). These non-MHC signals did not surpass genome-wide significance (*P* < 5×10^−8^): it is therefore unlikely that these variants represent genuine signals and more likely they reflect false positives due to statistical noise and/or population stratification. Test statistics were not inflated (λ_GC_< 1 in both SAS & AFR in the primary analysis), indicating adequate control for population stratification and other sources of bias. We observed striking consistency between statistical approaches (**supplementary figures 13-15**), with the mixed effects model used in the primary analysis yielding more conservative test statistics as expected due to the explicit adjustment for population structure (λ_GC_ 1.08 for SAS and 1.10 for AFR in the fixed effects analysis). The lack of association signals surpassing genome-wide significance (*P* < 5×10^−8^) was expected from statistical power considerations (**supplementary figure 16**). In the SAS GWAS, the MHC association signal lay within the class II HLA region (lead variant chr6:32600515:G:A, AF_MS_ = 0.42, AF_Controls_ = 0.30, OR 1.84, 95% CI 1.42 – 2.39, *P=*4.6×10^−6^, nearest gene *HLA-DRB1*), whereas in the AFR GWAS the strongest association lay within the class I HLA region (lead variant chr6:29919337:A:G, AF_MS_ = 0.26, AF_Controls_ = 0.15, OR 2.24, 95% CI 1.52 – 3.29, *P=*4.3×10^−5^, nearest gene *HLA-A*).

**Figure 3:**
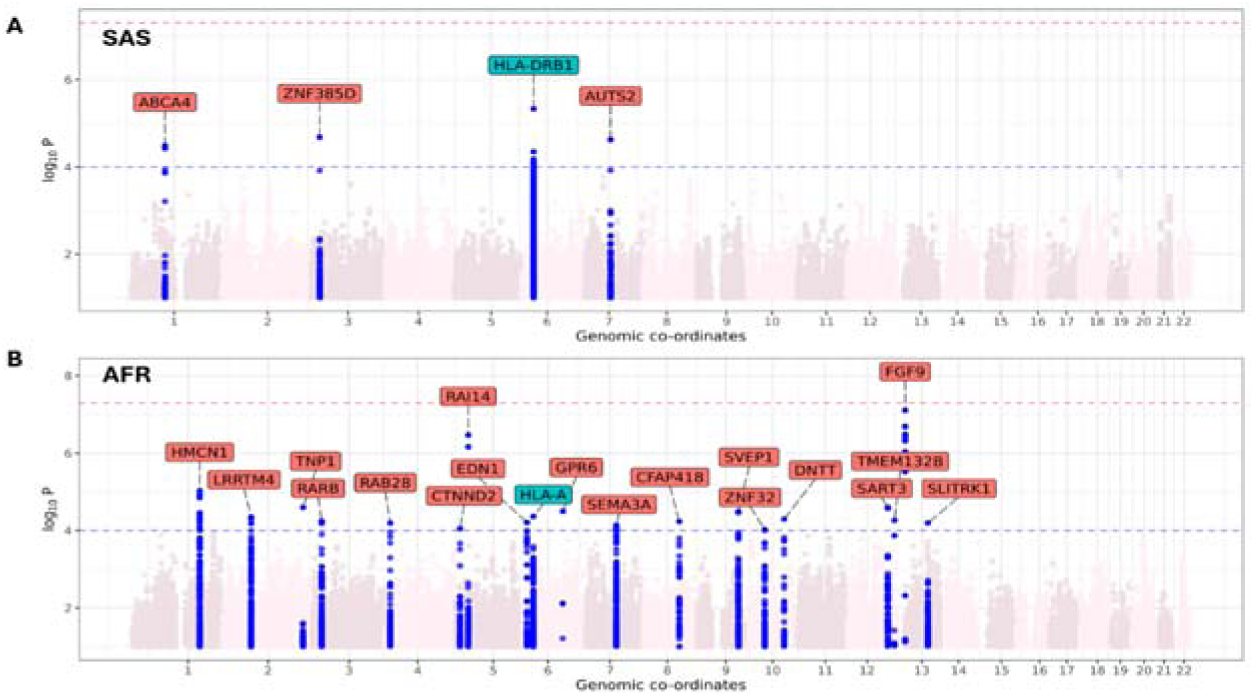
Genetic analysis of MS susceptibility in South Asian (SAS) and African (AFR) ancestries. A: Manhattan plot showing GWAS of MS risk in SAS genetic ancestry. Highlighted SNPs in blue reflect suggestive associations (*P* < 1×10^−4^). SNPs are annotated with the nearest gene. The nearest gene is labelled in blue if the variant was located within +/−1MB of a suggestive signal (*P* < 1×10^−5^) in the IMSGC European-ancestry discovery-stage GWAS, and in red if there were no nearby suggestive signals. B: as per A, but for AFR genetic ancestry.

**Table 2:**
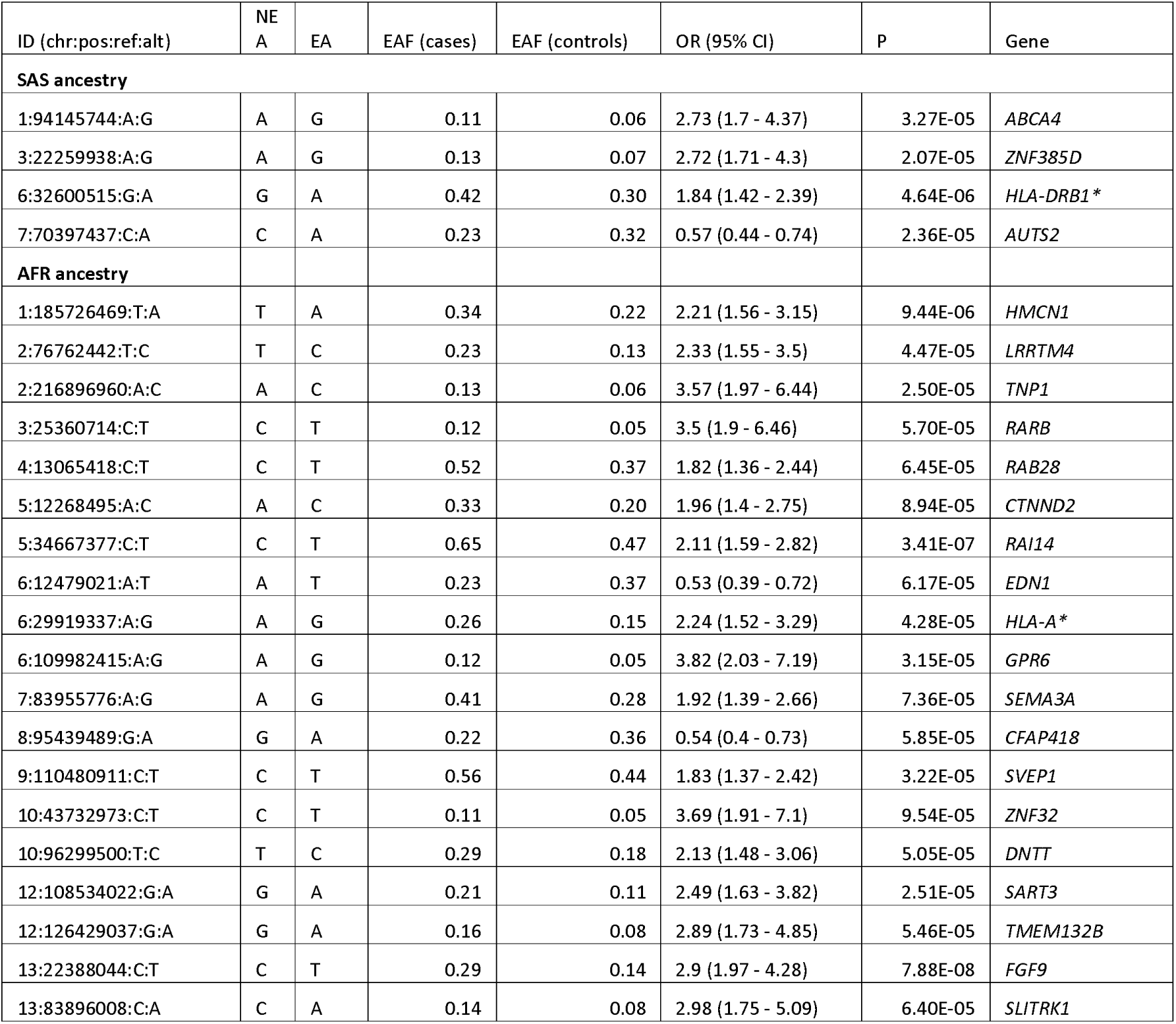
Suggestive (P < 1×10^−4^) genetic associations with MS risk in South Asian and African-ancestry MS cases. Columns show the SNP ID, Non-effect allele (NEA), effect allele (EA), the frequency of the effect allele (EAF) in cases and controls, the Odds Ratio (OR) with 95% confidence interval (CI), orientated to the effect allele, the P value for association, and the nearest gene.

| ID (chr:pos:ref:alt) | NE<br>A | EA | EAF (cases) | EAF (controls) | OR (95% CI) | P | Gene |
| --- | --- | --- | --- | --- | --- | --- | --- |
| **SAS ancestry** |  |  |  |  |  |  |  |
| 1:94145744:A:G | A | G | 0.11 | 0.06 | 2.73 (1.7 - 4.37) | 3.27E-05 | *ABCA4* |
| 3:22259938:A:G | A | G | 0.13 | 0.07 | 2.72 (1.71 - 4.3) | 2.07E-05 | *ZNF385D* |
| 6:32600515:G:A | G | A | 0.42 | 0.30 | 1.84 (1.42 - 2.39) | 4.64E-06 | *HLA-DRB1\** |
| 7:70397437:C:A | C | A | 0.23 | 0.32 | 0.57 (0.44 - 0.74) | 2.36E-05 | *AUTS2* |
| **AFR ancestry** |  |  |  |  |  |  |  |
| 1:185726469:T:A | T | A | 0.34 | 0.22 | 2.21 (1.56 - 3.15) | 9.44E-06 | *HMCN1* |
| 2:76762442:T:C | T | C | 0.23 | 0.13 | 2.33 (1.55 - 3.5) | 4.47E-05 | *LRRTM4* |
| 2:216896960:A:C | A | C | 0.13 | 0.06 | 3.57 (1.97 - 6.44) | 2.50E-05 | *TNP1* |
| 3:25360714:C:T | C | T | 0.12 | 0.05 | 3.5 (1.9 - 6.46) | 5.70E-05 | *RARB* |
| 4:13065418:C:T | C | T | 0.52 | 0.37 | 1.82 (1.36 - 2.44) | 6.45E-05 | *RAB28* |
| 5:12268495:A:C | A | C | 0.33 | 0.20 | 1.96 (1.4 - 2.75) | 8.94E-05 | *CTNND2* |
| 5:34667377:C:T | C | T | 0.65 | 0.47 | 2.11 (1.59 - 2.82) | 3.41E-07 | *RAI14* |
| 6:12479021:A:T | A | T | 0.23 | 0.37 | 0.53 (0.39 - 0.72) | 6.17E-05 | *EDN1* |
| 6:29919337:A:G | A | G | 0.26 | 0.15 | 2.24 (1.52 - 3.29) | 4.28E-05 | *HLA-A\** |
| 6:109982415:A:G | A | G | 0.12 | 0.05 | 3.82 (2.03 - 7.19) | 3.15E-05 | *GPR6* |
| 7:83955776:A:G | A | G | 0.41 | 0.28 | 1.92 (1.39 - 2.66) | 7.36E-05 | *SEMA3A* |
| 8:95439489:G:A | G | A | 0.22 | 0.36 | 0.54 (0.4 - 0.73) | 5.85E-05 | *CFAP418* |
| 9:110480911:C:T | C | T | 0.56 | 0.44 | 1.83 (1.37 - 2.42) | 3.22E-05 | *SVEP1* |
| 10:43732973:C:T | C | T | 0.11 | 0.05 | 3.69 (1.91 - 7.1) | 9.54E-05 | *ZNF32* |
| 10:96299500:T:C | T | C | 0.29 | 0.18 | 2.13 (1.48 - 3.06) | 5.05E-05 | *DNTT* |
| 12:108534022:G:A | G | A | 0.21 | 0.11 | 2.49 (1.63 - 3.82) | 2.51E-05 | *SART3* |
| 12:126429037:G:A | G | A | 0.16 | 0.08 | 2.89 (1.73 - 4.85) | 5.46E-05 | *TMEM132B* |
| 13:22388044:C:T | C | T | 0.29 | 0.14 | 2.9 (1.97 - 4.28) | 7.88E-08 | *FGF9* |
| 13:83896008:C:A | C | A | 0.14 | 0.08 | 2.98 (1.75 - 5.09) | 6.40E-05 | *SLITRK1* |

### Consistency of European susceptibility alleles across ancestries

To determine whether European-ancestry MS susceptibility alleles exert a similar effect across ancestries, we defined 164 independent European-ancestry signals from published GWAS^1^ and examined their impact in the SAS and AFR cohorts.

In general, European susceptibility alleles were over-represented in MS cases of both SAS and AFR ancestry (**figure 4, supplementary table 3**). This consistency was more pronounced for the SAS cohort (104 / 154 SNPs showing a concordant effect direction; one-tailed binomial *P* = 8.1×10^−6^, Spearman’s ρ = 0.31, Spearman’s *P* = 8.9×10^−5^) than the AFR cohort, in which there was a trend towards over-representation of European susceptibility alleles, but this enrichment did not achieve statistical significance (80 / 152 SNPs showing a concordant effect direction; one-tailed binomial *P* = 0.3, Spearman’s ρ = 0.1, Spearman’s *P* = 0.3).

**Figure 4:**
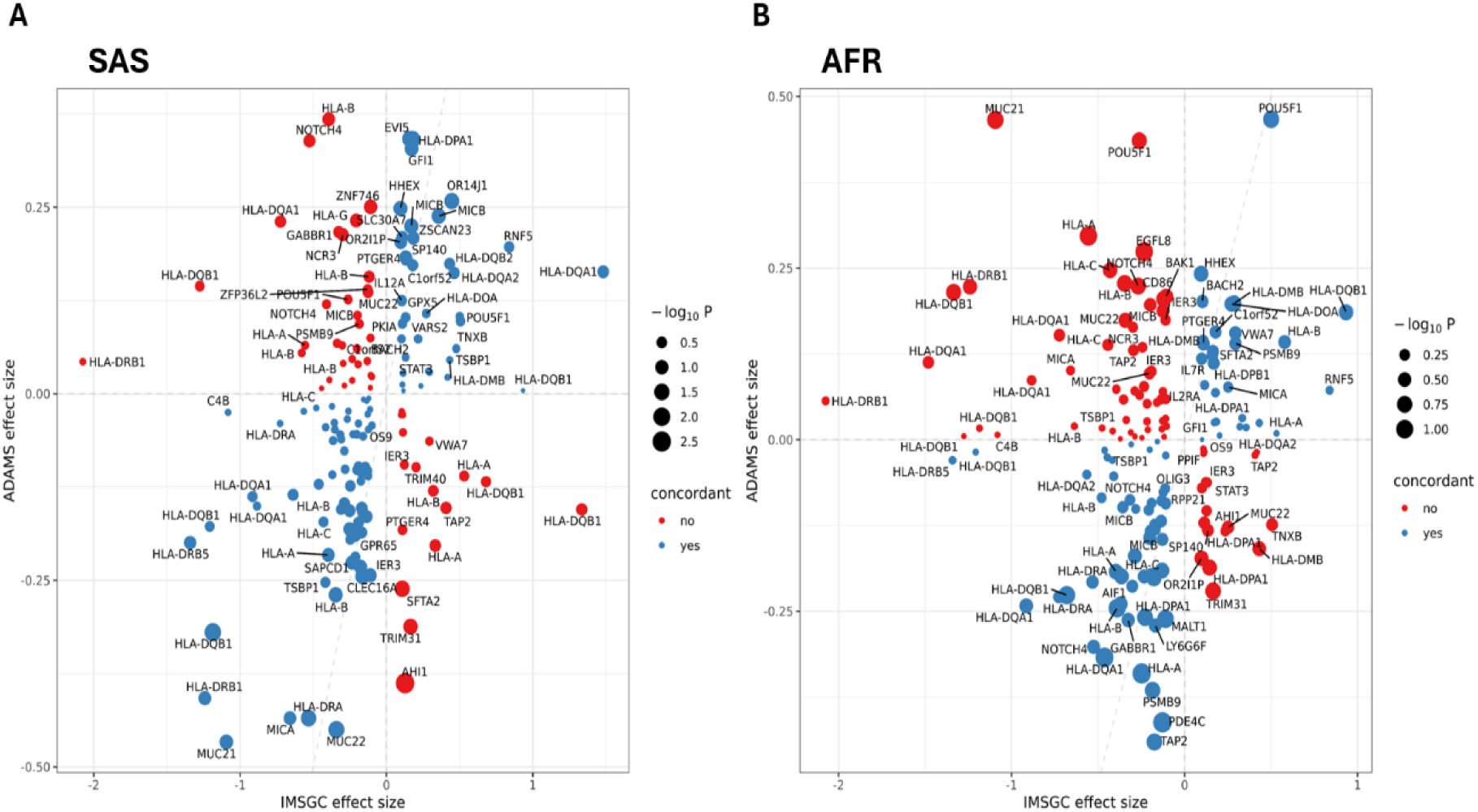
Consistency of European-ancestry MS risk alleles in South Asian (SAS) and African (AFR) ancestries. A: Beta-beta plot showing the effect sizes for genome-wide significant independent risk variants for MS derived from the 2019 IMSGC GWAS discovery-phase summary statistics. Each dot represents a genetic variant and is annotated with the nearest gene (some labels are omitted for clarity). The x-axis shows the effect size (beta, i.e. log odds ratio) of the variant in the IMSGC European-ancestry study. The y-axis shows the effect size in our South Asian ancestry cohort. The size of the points represents the P value in our cohort, i.e. larger points indicate more precisely estimated effects. Points are coloured according to their concordance across the ancestries, i.e. whether the effect direction (risk-increasing or decreasing) was the same in the South Asian cohort as in the original study. B: as per A, but for AFR genetic ancestry.

As the MHC locus is highly polymorphic across ancestries and contains multiple independent signals, the inclusion of this region dominates estimates of susceptibility allele concordance. However, excluding European alleles within the extended MHC locus did not appreciably alter our observation: the consistency of risk alleles remained strong in the SAS cohort (one-tailed binomial *P* = 0.007), and continued to trend in the same direction in the AFR cohort (one-tailed binomial *P* = 0.14). Importantly, the absolute correlation coefficients were of moderate magnitude only (ρ < 0.5), suggesting that European susceptibility alleles exert broadly similar effects across ancestries but that this correlation is imperfect, consistent with previous work^8,10,15,16,57^. This observation could be explained by ancestral differences (i.e. differences in the marginal SNP effects due to ancestral variation in Linkage Disequilibrium and allele frequency) or due to statistical imprecision of the effect estimates in our cohort, which may cause chance fluctuation of test statistics across the null. To explore the latter possibility, we repeated the analysis restricting to SNPs with a *P* value of <0.5. When restricting to these variants, we observed strong evidence of consistency in both the AFR (45/ 69 SNPs concordant, *P* = 0.008) and SAS (57 / 79 SNPs, *P* = 5.1×10^−5^) cohorts. None of the European-ancestry variants achieved suggestive significance (*P* < 1×10^−4^) in either cohort (**supplementary table 3**).

### Classical HLA allelic associations in MS across ancestries

Given the SNP-level associations with MS susceptibility at the MHC locus in both SAS and AFR ancestry, we sought to determine whether classical HLA allelic associations drive these associations. To do so we imputed classical HLA alleles at six genes (*HLA-A*, *HLA-B*, *HLA-C*, *HLA-DPB1*, *HLA-DRB1*, *HLA-DQB1*) using a multi-ancestry imputation panel, and validated these calls using two different imputation panels and two separate methods of imputation. We found several suggestive associations between specific classical HLA alleles and MS susceptibility in both ancestries (**figure 5, supplementary table 4**). In the SAS cohort, we observed eight associations at a False Discovery Rate of <10%: risk-increasing effects of *HLA-DPB1*10:01* (OR 3.1, 95% CI 1.9 – 5.2), *HLA-B*37:01* (OR 2.2, 95% CI 1.5 – 3.4), *HLA-A*26:01* (OR 1.8, 95% CI 1.3 – 2.5), *HLA-DRB1*15:01* (OR 1.7, 95% CI 1.2 – 2.3), *HLA-A*23:01* (OR 2.7, 95% CI 1.4 – 5.2), and *HLA-DRB1*04:01* (OR 2.5, 95% CI 1.3 – 4.8), and protective effects of *HLA-DRB1*13:01* (OR 0.4, 95% CI 0.2 – 0.7) and *HLA-DQB1*06:03* (OR 0.4, 95% CI 0.2 – 0.7). In the AFR cohort, the risk-increasing impact of *HLA-A*66:01* (OR 3.5, 95% CI 1.7 – 7) was the only signal surpassing a False Discovery Rate of 10%, however we observed suggestive (*P* < 0.05) associations with *HLA-DRB1*15:01* (OR 2.3, 95% CI 1.2 – 4.3, *P* = 0.01), *DRB1*01:02* (OR 1.7, 95% CI 1.1 – 2.8), *DRB1*11:02* (OR 0.2, 95% CI, and *HLA-B*57:03* (OR 1.9, 95% CI 1.1 – 3.1). Many of these alleles are rare (MAF between 1% and 5%) and so prone to statistical noise – of the associations reported, the only alleles present at MAF >=5% in both cases and controls were *DRB1*15:01* and *A*26:01*.

**Figure 5:**
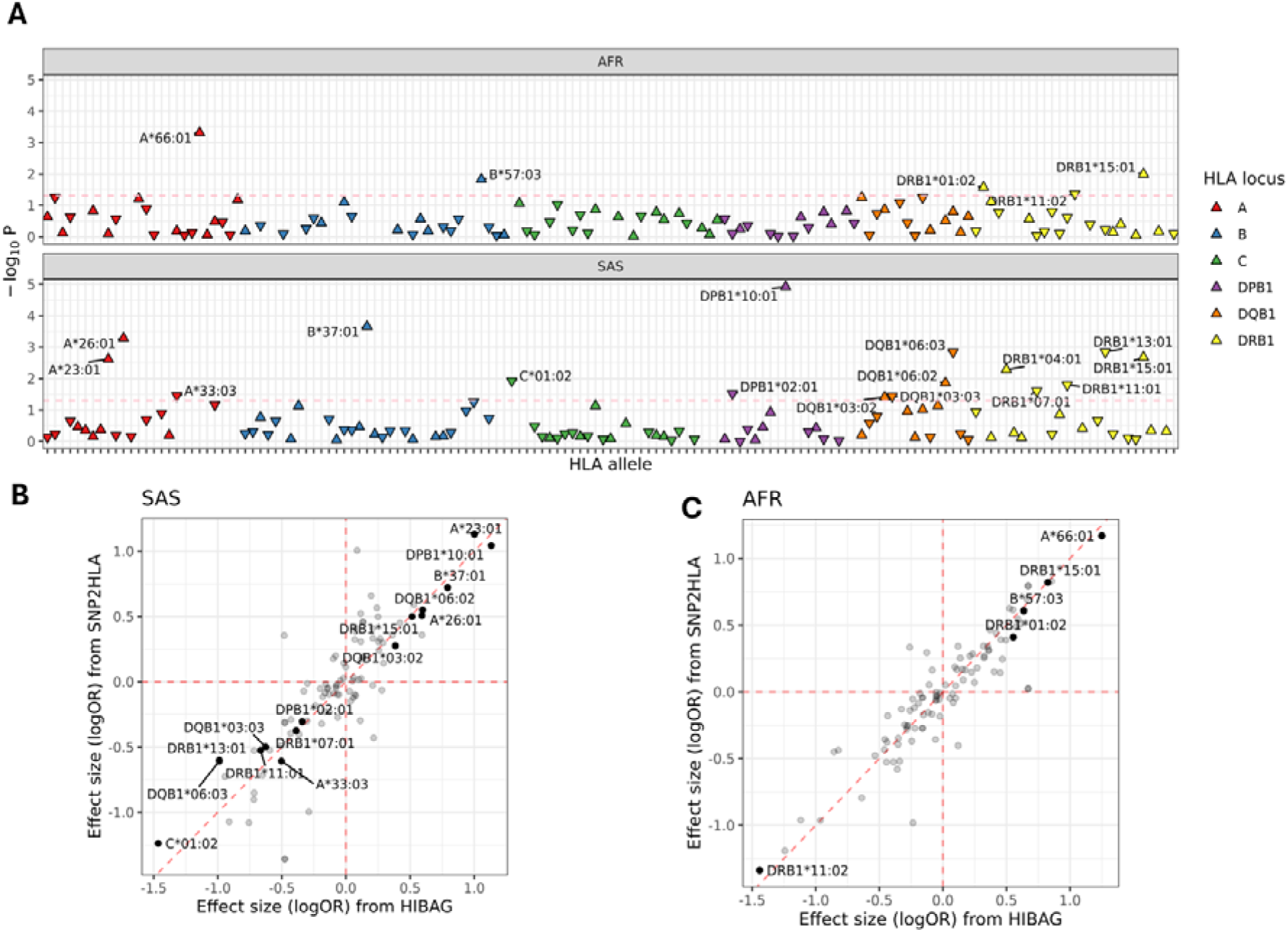
HLA classical allelic associations with MS in South Asian (SAS) and African (AFR) ancestries. A: Panels show the results from association testing between classical HLA alleles and MS risk in AFR (top) and SAS (bottom) ancestries. Only alleles with an overall allele frequency of >1% were tested in each ancestry. The x-axis indicates the allele. The y-axis indicates the strength of the association. The horizontal dashed line indicates nominal significance, i.e. *P* < 0.05. P values were derived from multivariable logistic regression models adjusted for sex and the first ten PCs. HLA alleles were treated using an additive genetic model, i.e. the exposure variable was ‘0’, ‘1’, or ‘2’ for each allele. Each point indicates an allele, the colours indicate the locus. Points are orientated in their effect direction, i.e. ‘risk-increasing’ alleles are shown as upward triangles, ‘protective’ alleles are shown as downward triangles. Only alleles showing nominal association at *P* < 0.05 are labelled with the allele name for clarity. HLA alleles were imputed from SNP data using HIBAG. B: comparison of effect sizes (log Odds Ratio for MS given HLA allelic dosage) derived from HIBAG (the primary analysis, x-axis) and SNP2HLA (y-axis). The diagonal red line indicates the null, i.e. identical effect sizes. Dots are greyed out if they did not achieve nominal significance (*P* < 0.05) in the primary analysis. Only allele labels for the nominally-associated alleles are shown. This panel shows the SAS-ancestry results. C: as per B, but for the African-ancestry study.

To methodologically validate these associations we repeated these analyses using an alternative imputation framework (SNP2HLA). Effect sizes, allele frequencies, and statistical significance were consistent between methods **(figure 5B & C, supplementary table 4).** All the associations achieving an FDR < 10% using the HIBAG method (SAS: *DPB1*10:01*, *B*37:01*, *A*26:01*, *DRB1*13:01*, *DQB1*06:03*, *DRB1*15:01* and *A*23:01*; AFR: *A*66:01*) were validated at *P* < 0.1 using SNP2HLA (all except *DRB1*13:01* had *P* < 0.05). *HLA-DRB1*04:01* was not tested in the sensitivity analysis as the imputed allele frequency was substantially lower (MAF < 1%) with SNP2HLA.

To further confirm the validity of our imputation, we created a visualisation tool (https://benjacobs.shinyapps.io/ukb_hla_freq_browser/) to explore the frequency of classically imputed HLA alleles in the entire UK Biobank dataset across ancestries, taking as its input the alleles imputed by the UK Biobank using an alternative algorithm (HLA*IMP). We confirmed similar control frequencies using our approach compared with the HLA*IMP approach in the entire UK Biobank dataset, e.g. for the African-ancestry signal *A*66:01* we see a control AF of 0.012 vs 0.015 in UKB, and for the lead South Asian-ancestry signal we see similar concordance (*DPB1*10:01* AF 0.018 vs 0.013).

We performed stepwise conditional analysis to determine whether these marginal associations were independent of one another, and whether there were further allelic associations masked by LD (**supplementary table 5**). We chose a *P* value threshold of <0.01 for the stepwise conditioning (i.e. we continued the conditional analysis until no further associations persisted at *P* < 0.01). In the SAS cohort, this analysis confirmed independent effects of *DPB1*10:01*, *A*26:01*, *B*37:01*, *A*23:01*, *DRB1*15:01*, and *DQB1*03:02*. In the AFR cohort, conditioning on *A*66:01* yielded a suggestive association with *A*29:02*, and subsequently with *B*57:03* (**supplementary table 5**).

### Consistency and differences at the MHC locus across ancestries

Of the associations reported in European ancestry cases, most alleles showed at least nominal evidence of replication in persons of South Asian and African ancestry where the allele was common enough and sufficiently well-imputed to test (**table 3**). All European-ancestry risk alleles showed concordant effect directions in South Asian and African ancestries (**table 3**). Importantly, while these alleles likely exert a similar effect if present in people of African or South Asian ancestry, their frequency is much lower, so the population-level relevance of these alleles is less than that of European populations. We estimated the population attributable fraction (PAF) for *HLA-DRB1*15:01*, i.e. the overall burden of MS prevalence explained by the presence of this allele in the population. Assuming a dominant model (i.e. comparing the presence of one or two *DRB1*15:01* alleles vs no *DRB1*15:01* alleles), an approximate and conservative effect size of Relative Risk ∼ 3.9, and prevalence of at least one *DRB1*15:01* allele in ∼50% of cases (consistent with UK Biobank European-ancestry MS cases^58^), the PAF of *HLA-DRB1*15:01* in European-ancestry MS is 43.7%. In contrast, our findings suggest that the PAF of *HLA-DRB1*15:01* in people of South Asian ancestry is 9.8% (OR 1.7, AF_Case_ 0.13), and 4.5% in people of African-ancestry (OR 2.3, AF_Case_ 0.05). We obtained similar results using Miettinen’s approximation (PAF_EUR_ 60.1%, PAF_SAS_ 9.5%, PAF_AFR_ 5.4%).

**Table 3:** effect estimates for classical HLA alleles reported in European-ancestry MS cases, and in the ADAMS cohort (SAS & AFR ancestry). ‘NA’ indicates that the allele was either too rare (AF < 1%) or imputed with too little accuracy in that ancestry to test. Columns show the HLA allele, allele frequency (AF) in cases and controls, and Odds Ratios with 95% Confidence Intervals for the European-ancestry IMSGC study, and our SAS and AFR-ancestry cohorts.

| HLA allele | EUR (Moutsianas *et al*) |  | SAS |  |  |  | AFR |  |  |  |
| --- | --- | --- | --- | --- | --- | --- | --- | --- | --- | --- |
|  | AF (Cont) | OR (95% CI) | OR (95% CI) | P | AF (Cont) | AF (Case) | OR (95% CI) | P | AF (Cont) | AF (Case) |
| *DRB1\*15:01* | 0.143 | 3.92 (3.74–4.12) | 1.67 (1.2 - 2.3) | 0.002 | 0.085 | 0.126 | 2.28 (1.2 - 4.3) | 0.010 | 0.018 | 0.049 |
| *A\*02:01* | 0.276 | 0.67 (0.64–0.70) | 0.88 (0.5 - 1.4) | 0.587 | 0.060 | 0.054 | 0.59 (0.3 - 1) | 0.056 | 0.098 | 0.062 |
| *DRB1\*03:01* | 0.143 | 1.16 (1.10–1.22) | 1.05 (0.8 - 1.4) | 0.750 | 0.114 | 0.134 | 1.52 (1 - 2.4) | 0.076 | 0.063 | 0.093 |
| *DRB1\*13:03* | 0.009 | 2.62 (2.32–2.96) | NA | NA | NA | NA | 1.28 (0.7 - 2.3) | 0.399 | 0.047 | 0.058 |
| *DRB1\*08:01* | 0.020 | 1.55 (1.42–1.69) | NA | NA | NA | NA | NA | NA | NA | NA |
| *B\*44:02* | 0.114 | 0.78 (0.74–0.83) | 0.65 (0.2 - 2) | 0.459 | 0.011 | 0.009 | NA | NA | NA | NA |
| *B\*38:01* | 0.011 | 0.48 (0.42–0.56) | NA | NA | NA | NA | NA | NA | NA | NA |
| *B\*55:01* | 0.018 | 0.63 (0.55–0.73) | 0.39 (0.1 - 1.2) | 0.106 | 0.021 | 0.009 | NA | NA | NA | NA |
| *DQA1\*01:01* | 0.144 | 0.65 (0.59–0.72) | NA | NA | NA | NA | NA | NA | NA | NA |
| *DQB1\*03:02* | 0.105 | 1.30 (1.23–1.37) | 1.47 (1 - 2.1) | 0.039 | 0.074 | 0.097 | 1.64 (0.9 - 3.2) | 0.137 | 0.028 | 0.044 |
| *DQB1\*03:01* | 0.187 | 0.60 (0.52–0.69) | 0.79 (0.6 - 1.1) | 0.158 | 0.128 | 0.111 | 0.68 (0.4 - 1.2) | 0.183 | 0.080 | 0.058 |

Some of the associations we report (at FDR <10%) have been reported previously in European-ancestry MS^1,6,52^, i.e. the risk-increasing effects of *HLA-DRB1*15:01*, *HLA-B*37:01*; and the protective effects of *HLA-DRB1*13:01* and *DQB1*06:03*. The other associations in South Asian (*HLA-DPB1*10:01*, *HLA-A*26:01*, *HLA-A*23:01*) and African-ancestry (*A*66:01*) cohorts have not been previously reported. This apparent novelty may reflect either type I error (i.e. false positive results) or genuinely ancestry-specific associations. In UK Biobank, *HLA-A*66:01* is enriched in the African-ancestry participants vs the European-ancestry participants (AF_AFR_ 0.02 vs AF_EUR_ 0.003, fold-change 5.2x), and *HLA-A*26:01* is weakly enriched in the South Asian ancestry participants (AF_SAS_ 0.06 vs AF_EUR_ 0.02, fold-change 2.7x). Neither *DPB1*10:01* nor *A*23:01* show enrichment in the South Asian participants.

### Genetic risk prediction in non-European ancestries

To further quantify the degree of genetic overlap between the architecture of MS risk in European and non-European ancestry populations, we derived polygenic risk scores (PRS) from published European-ancestry summary statistics and applied these scores to the participants of South Asian and African ancestry in our cohort. When generating a genetic risk score in one ancestry and applying it to another, it is expected that there will be some drop-off in performance (roughly related to the genetic distance between populations)^59^ due to ancestral differences in allele frequency and LD. However, assuming at least some genetic overlap in the loci involved, these scores should still have some discriminative power in non-European ancestries.

European-derived PRS distinguished MS cases from controls at a population level across ancestries (**figure 6**). In keeping with the hypothesis that there is at least partial overlap in the genetic architecture of MS susceptibility across ancestries, the PRS was statistically predictive of MS risk in both SAS and AFR ancestries, but consistently performed poorer than published European-ancestry estimates (both in terms of empirical *P* value fit and the liability explained by the PRS, **figure 6**). The best-performing PRS explained 1.6% of the liability to MS among SAS-ancestry participants (N_SNPs_ = 174, empirical *P* = 1.0×10^−4^) and 0.5% among AFR-ancestry participants (N_SNPs_ = 298, empirical *P* = 0.08). For comparison, the European-ancestry estimate for the liability explained by a PRS from our previous work in European-ancestry MS cases in UK Biobank (using a far greater number of controls and slightly different set of genetic variants) was 4.3%^16^.

**Figure 6:**
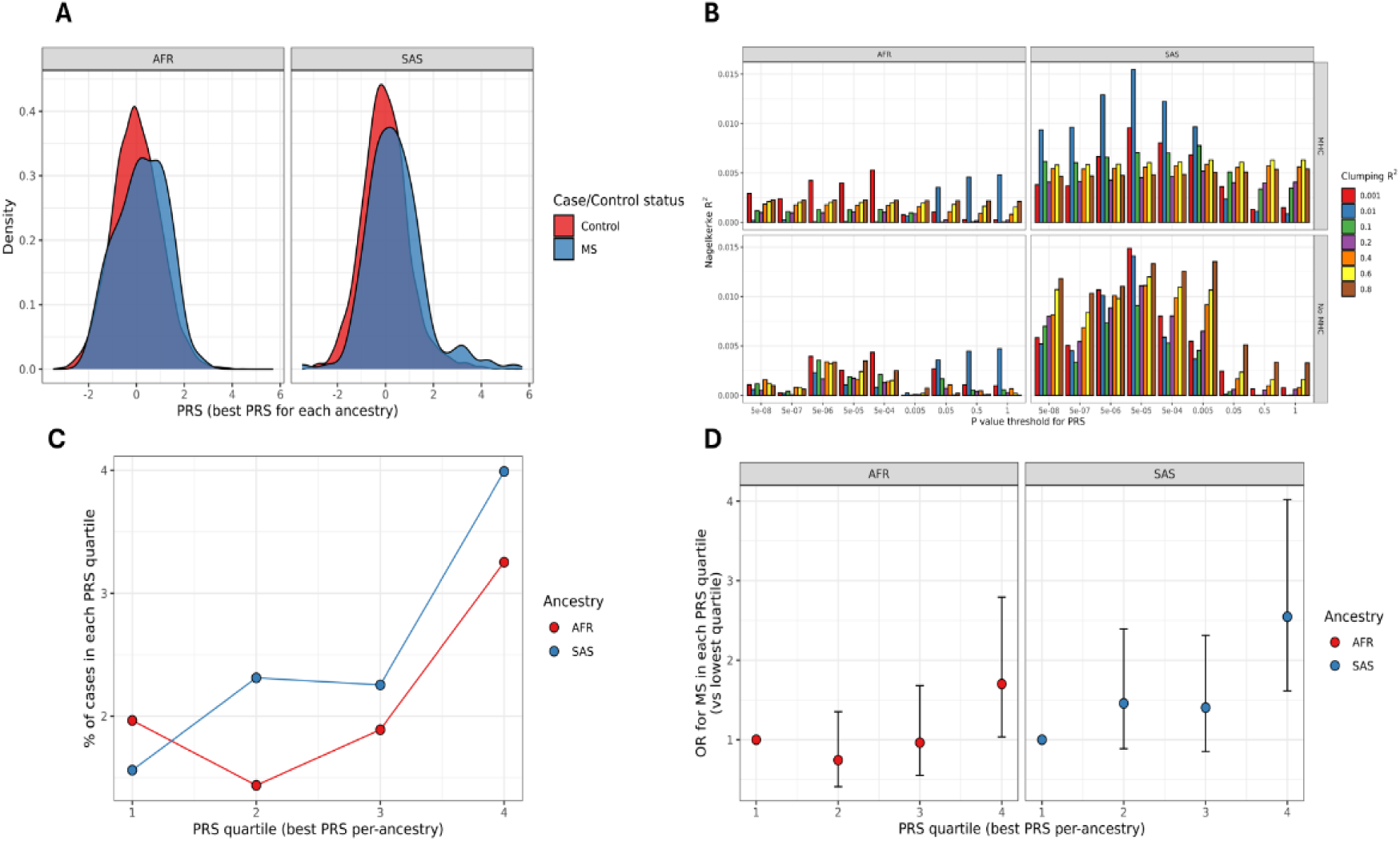
European-derived polygenic risk score performance in MS cases of South Asian (SAS) and African (AFR) ancestries. A: Density plots showing the distribution of PRS among cases & controls for SAS and AFR cohorts. The plots demonstrate a tendency for cases to have higher PRS than controls but sizeable overlap in the distributions, in keeping with expectation from PRS studies in European-ancestry MS. B: Barplots showing the discriminative performance – Nagelkerke’s pseudo-R^2^ – in SAS and AFR cohorts across a range of PRS tuning parameters. The y-axis corresponds roughly to the liability explained by the PRS alone, after adjustment for covariates (sex, and PCs 1-10). The x-axis shows the *P* value threshold used to select variants for the PRS (i.e. for a P value threshold of 0.001, only SNPs showing association with MS in the IMSGC study at P < 0.001 were included for the PRS). The colours of the bars indicate the clumping R^2^ parameter used to define independent SNPs for the PRS. The MHC vs non-MHC panels indicate whether the MHC region was included (‘MHC’) or excluded (‘No MHC’) from the PRS. C: calibration plots showing, for the best PRS in each ancestry, the absolute observed prevalence of cases in each PRS quartile, indicating a broadly monotonic relationship with more MS among the top quartile. D: Dot plot showing the odds ratio derived from logistic regression analysis for MS given the PRS quartile, compared with the lowest quartile (reference, OR = 1). Regression models were adjusted for sex & PCs 1-10. Error bars indicate the 95% confidence intervals.

We quantified PRS calibration by calculating the proportion of MS cases within each quartile of the best-performing PRS in each ancestry. Broadly speaking, there was a monotonic relationship between PRS quartile and the proportion of cases in all ancestries (**figure 6**). Both the magnitude of this effect and the statistical strength of evidence were more pronounced in the SAS ancestry cohort (**figure 6**). These findings are consistent with strong but imperfect overlap of European risk alleles with South Asian and African-ancestry MS.

## Discussion

Here we present a genome-wide association study of Multiple Sclerosis risk in a UK cohort of South Asian and African ancestry, complementing and extending previous consortium-level efforts in European-ancestry MS (N_MS_ ∼50,000), and building on studies in other non-European ancestral groups, including African Americans with MS (N_MS_ ∼1400)^10^, in India (N_MS_ ∼ 270)^12^, in Hispanic Americans (N_MS_ = 1298)^8^, and in Japan (N ∼ 145)^13^.

By contrasting the genetic profiles of our diverse MS cohort with ancestrally-similar controls from UK Biobank, we saw the anticipated evidence for association between MHC variants and MS susceptibility. Although we found no evidence for novel association, we saw overwhelming evidence that genetic risk factors identified in European populations also influence risk in other populations. The concordance of risk alleles across populations argues for shared disease mechanisms. Studies involving much greater numbers of cases will be needed to leverage genetic diversity to power novel discoveries, i.e. to enhance the fine-mapping of established associations, and to identify risk variants that are rare in Europeans but more common in other populations.

Fine-mapping of the MHC signal revealed points of convergence with the European-ancestry association patterns within this locus. For example, the major European risk alleles – including the strongest genetic risk factor for MS, *HLA-DRB1*15:01* - showed highly concordant effects in the South Asian and African-ancestry cohorts. The lower frequency of *HLA-DRB1*15:01* in South Asian and African ancestral groups translates into a lower population relevance of this allele.

In the largest published analysis of the HLA association in South Asian individuals with MS, the strongest signal was for *HLA-DRB1*15:01*, followed by a conditionally independent effect of *HLA-DRB1*03:01*^11^. There was a nominal effect observed for *HLA-DQA1*05:01* which dissipated on adjustment for *DRB1*15:01*, suggesting that a *DRB1*15:01*-containing haplotype drove this association. No associations were observed for *DQB1* alleles. Our analysis replicates and extends these findings. In our cohort of South Asian ancestry, we found a strong HLA allelic association with the *HLA-DRB1*15:01* allele, but the strongest association was for the *DPB1* allele *DPB1*10:01*. While this allele is shared globally, it may tag an effect of haplotypes containing *DRB1*10:01* and *DQB1*05:01*, which is strongly enriched in South Asian populations. The largest analysis of the HLA in African ancestry cases with MS was performed in an African American cohort – a heterogeneous population with two-way European-African admixture – and so is slightly different from the cohort presented here^9^. This study demonstrated independent risk-increasing effects of *DRB1*15:01*, *DRB1*03:03*, and *DRB1*04:05*, with protective effects of *DRB1*11:01*, *DRB1*04:04* and *A*02:01*, and no effect of *DQB1* alleles. The findings from our cohort broadly replicate and extend these findings – we find a concordant effect direction (albeit not significant at FDR < 10%) for *HLA-DRB1*15:01*. In our cohort, the strongest effect at this locus was the risk-increasing effect of *HLA-A*66:01*, which is strongly enriched in African-ancestry populations and so may similarly reflect a population-specific signal. In summary, our analyses have highlighted concordant effect directions for European-ancestry HLA risk alleles in African and South Asian MS, and suggested some additional alleles which have not been previously reported and require external validation in larger cohorts.

A limitation of this work is the reliance on existing HLA imputation panels to infer HLA alleles rather than using gold-standard methods (PCR or sequencing). This concern is particularly relevant given the highly polymorphic nature of the MHC and the relatively limited genetic diversity present in the reference panels used. We aim to obviate some of these concerns by cross-validating HLA allele calls with an alternative method (and an alternative reference panel), and we are reassured by the consistency of findings across the two methods. Increasing the availability of multi-ancestry HLA imputation panels will help to clarify the contribution of the MHC region in the future and help define both normal and disease-associated genetic variation at this complex locus across ancestries.

The other limitation of this work is using cases and controls from largely distinct cohorts. This can introduce batch effect due to differences in allele frequency purely related to technical factors (such as blood vs saliva-derived DNA, genotyping chip, and imputation panel), differences in the ancestral composition of the cohorts, and other unbalanced confounders such as age, gender, and demographic factors. We have used a stringent quality control procedure to filter out variants at risk of bias, including removing variants that differ in frequency between MS cases from UK Biobank and our cohort and performing re-imputation using a combined dataset with this filtered set of variants. Regarding ancestral differences in the cohorts, we use a stringent, multi-step approach to estimating genetic ancestry and excluding PCA outliers to minimise the risk of population stratification. The use of the mixed models implemented in REGENIE further accounts for population structure by regressing out the impact of genome-wide variation. Our quality control pipeline is deliberately conservative to attempt to minimise type 1 error, as evidenced by the genome-wide deflation of test statistics and the lack of genome-wide significant associations in either cohort. Importantly, none of the results we highlight achieved a stringent genome-wide significance threshold of *P* < 5×10^−8^, and so these results should be interpreted as a preliminary effort which needs to be expanded and replicated with larger multi-ancestry genetic cohorts.

The major weakness of this study is the sample size. Despite being the biggest UK multi-ancestry genetic study of MS to date, this study remains underpowered to detect novel effects. Other limitations of our approach include our use of very strict ancestry definitions, resulting in several cases being excluded as PCA outliers, and the reliance on self-reported MS.

In summary, we have undertaken a genetic analysis of Multiple Sclerosis susceptibility in people of South Asian and African ancestry living in the UK, demonstrating the universal importance of the MHC locus in MS risk, and the striking correlation in genetic architecture across ancestries. Despite being a major undertaking, this study is very small for a genetic study of a complex disease. Hopefully, this represents an early initial step towards a well-powered, multi-ancestry GWAS effort, similar to the successful efforts in other complex traits and diseases^19–22^. With appropriate sample sizes (in the tens of thousands), multi-ancestry GWAS has the potential to discover novel risk variants for MS, fine-map existing associations to identify causal signals, leverage these causal insights to find new and improved drug targets, and enhance genetic prediction of MS. This will only be possible by expanding this cohort and combining these data with international colleagues in the next consortium-level meta-analysis of MS risk.

## Data and code availability

Code used to perform these analyses is provided on Github at https://github.com/benjacobs123456/ms_geno_diversity. Genetic association summary statistics will be made available via the GWAS catalogue on publication.

## Acknowledgements

We would like to acknowledge the generosity of all the people with MS who contributed to the study, the International Multiple Sclerosis Genetics Consortium (IMSGC) for provision of summary association statistics from the European-ancestry discovery GWAS, and all other contributors who provided software, code, or data used in this study.

## References

1. International Multiple Sclerosis Genetics Consortium. Multiple sclerosis genomic map implicates peripheral immune cells and microglia in susceptibility. Science 365, (2019).

2. International Multiple Sclerosis Genetics Consortium. Electronic address: & International Multiple Sclerosis Genetics Consortium. Low-Frequency and Rare-Coding Variation Contributes to Multiple Sclerosis Risk. Cell 178, 262 (2019).

3. Sawcer, S., Franklin, R. J. M. & Ban, M. Multiple sclerosis genetics. Lancet Neurol. 13, 700–709 (2014).

4. Analysis of immune-related loci identifies 48 new susceptibility variants for multiple sclerosis. Nat. Genet. 45, 1353–1360 (2013).

5. International Multiple Sclerosis Genetics Consortium et al. Genetic risk and a primary role for cell-mediated immune mechanisms in multiple sclerosis. Nature 476, 214–219 (2011).

6. Moutsianas, L. et al. Class II HLA interactions modulate genetic risk for multiple sclerosis. Nat. Genet. 47, 1107–1113 (2015).

7. Chi, C. et al. Admixture mapping reveals evidence of differential multiple sclerosis risk by genetic ancestry. PLoS Genet. 15, e1007808 (2019).

8. Beecham, A. H. et al. The genetic diversity of multiple sclerosis risk among Hispanic and African American populations living in the United States. Mult. Scler. 26, 1329–1339 (2020).

9. Isobe, N. et al. Genetic risk variants in African Americans with multiple sclerosis. Neurology 81, 219–227 (2013).

10. Isobe, N. et al. An ImmunoChip study of multiple sclerosis risk in African Americans. Brain 138, 1518–1530 (2015).

11. Pandit, L. et al. HLA associations in South Asian multiple sclerosis. Mult. Scler. 22, 19–24 (2016).

12. Pandit, L. et al. Evaluation of the established non-MHC multiple sclerosis loci in an Indian population. Mult. Scler. 17, 139–143 (2011).

13. Yoshimura, S. et al. Genetic and infectious profiles of Japanese multiple sclerosis patients. PLoS One 7, e48592 (2012).

14. Jacobs, B. M. et al. Towards a global view of multiple sclerosis genetics. Nat. Rev. Neurol. 18, 613–623 (2022).

15. Rivier, C. A. et al. Differential results of polygenic risk scoring for multiple sclerosis in European and African American populations. medRxiv 2024.06.11.24308714 (2024) doi:10.1101/2024.06.11.24308714.

16. Breedon, J. R. et al. Polygenic risk score prediction of multiple sclerosis in individuals of South Asian ancestry. Brain Communications 5, fcad041 (2023).

17. Duncan, L. et al. Analysis of polygenic risk score usage and performance in diverse human populations. Nat. Commun. 10, 3328 (2019).

18. Goris, A., Vandebergh, M., McCauley, J. L., Saarela, J. & Cotsapas, C. Genetics of multiple sclerosis: lessons from polygenicity. Lancet Neurol. 21, 830–842 (2022).

19. Graham, S. E. et al. The power of genetic diversity in genome-wide association studies of lipids. Nature 600, 675–679 (2021).

20. Chen, M.-H. et al. Trans-ethnic and Ancestry-Specific Blood-Cell Genetics in 746,667 Individuals from 5 Global Populations. Cell 182, 1198–1213.e14 (2020).

21. Vujkovic, M. et al. Discovery of 318 new risk loci for type 2 diabetes and related vascular outcomes among 1.4 million participants in a multi-ancestry meta-analysis. Nat. Genet. 52, 680–691 (2020).

22. Chen, J. et al. The trans-ancestral genomic architecture of glycemic traits. Nat. Genet. 53, 840–860 (2021).

23. Oksenberg, J. R. et al. Mapping multiple sclerosis susceptibility to the HLA-DR locus in African Americans. Am. J. Hum. Genet. 74, 160–167 (2004).

24. Hollenbach, J. A. & Oksenberg, J. R. The immunogenetics of multiple sclerosis: A comprehensive review. J. Autoimmun. 64, 13–25 (2015).

25. Beecham, A. H. et al. Ancestral risk modification for multiple sclerosis susceptibility detected across the Major Histocompatibility Complex in a multi-ethnic population. PLoS One 17, e0279132 (2022).

26. Steri, M. et al. Overexpression of the Cytokine BAFF and Autoimmunity Risk. N. Engl. J. Med. 376, 1615–1626 (2017).

27. Jacobs, B. M. et al. Cohort profile: ADAMS project: a genetic Association study in individuals from Diverse Ancestral backgrounds with Multiple Sclerosis based in the UK. BMJ Open 13, (2023).

28. Jacobs, B. M. et al. The relationship between ethnicity and multiple sclerosis characteristics in the United Kingdom: A UK MS Register study. Mult. Scler. 30, 1544–1555 (2024).

29. Lerede, A. et al. Patient-reported outcomes in multiple sclerosis: a prospective registry cohort study. Brain Commun. 5, fcad199 (2023).

30. Rodgers, W. J. et al. The impact of smoking cessation on multiple sclerosis disease progression. SSRN Electron. J. (2021) doi:10.2139/ssrn.3807268.

31. Ford, D. V. et al. The feasibility of collecting information from people with Multiple Sclerosis for the UK MS Register via a web portal: characterising a cohort of people with MS. BMC Med. Inform. Decis. Mak. 12, 73 (2012).

32. Kuri, A. et al. Evaluation of remote assessments for multiple sclerosis in an in-home setting. Mult. Scler. Relat. Disord. 54, 103125 (2021).

33. Hobart, J., Lamping, D., Fitzpatrick, R., Riazi, A. & Thompson, A. The Multiple Sclerosis Impact Scale (MSIS-29): a new patient-based outcome measure. Brain 124, 962–973 (2001).

34. Herdman, M. et al. Development and preliminary testing of the new five-level version of EQ-5D (EQ-5D-5L). Qual. Life Res. 20, 1727–1736 (2011).

35. Manouchehrinia, A. et al. Age Related Multiple Sclerosis Severity Score: Disability ranked by age. Mult. Scler. 23, 1938–1946 (2017).

36. Loh, P.-R. et al. Reference-based phasing using the Haplotype Reference Consortium panel. Nat. Genet. 48, 1443–1448 (2016).

37. McCarthy, S. et al. A reference panel of 64,976 haplotypes for genotype imputation. Nat. Genet. 48, 1279–1283 (2016).

38. Bycroft, C. et al. The UK Biobank resource with deep phenotyping and genomic data. Nature 562, 203–209 (2018).

39. Cavalli-Sforza, L. L. The Human Genome Diversity Project: past, present and future. Nat. Rev. Genet. 6, 333–340 (2005).

40. 1000 Genomes Project Consortium et al. A global reference for human genetic variation. Nature 526, 68–74 (2015).

41. Mbatchou, J. et al. Computationally efficient whole-genome regression for quantitative and binary traits. Nat. Genet. 53, 1097–1103 (2021).

42. Chang, C. C. et al. Second-generation PLINK: rising to the challenge of larger and richer datasets. Gigascience 4, 7 (2015).

43. McLaren, W. et al. The Ensembl Variant Effect Predictor. Genome Biol. 17, 122 (2016).

44. Karczewski, K. J. et al. The mutational constraint spectrum quantified from variation in 141,456 humans. Nature 581, 434–443 (2020).

45. Zheng, X. et al. HIBAG--HLA genotype imputation with attribute bagging. Pharmacogenomics J. 14, 192–200 (2014).

46. Zheng, X. Imputation-Based HLA Typing with SNPs in GWAS Studies. in HLA Typing: Methods and Protocols (ed. Boegel, S.) 163–176 (Springer New York, New York, NY, 2018).

47. Vince, N. et al. SNP-HLA Reference Consortium (SHLARC): HLA and SNP data sharing for promoting MHC-centric analyses in genomics. Genet. Epidemiol. 44, 733–740 (2020).

48. Silva, N. S. B., et al. 18th International HLA and Immunogenetics Workshop: Report on the SNP-HLA Reference Consortium (SHLARC) component. HLA 103, e15293 (2024).

49. Jia, X. et al. Imputing Amino Acid Polymorphisms in Human Leukocyte Antigenes. PLoS One 8, (2013).

50. Gonzalez-Galarza, F. F. et al. Allele frequency net database (AFND) 2020 update: gold-standard data classification, open access genotype data and new query tools. Nucleic Acids Res. 48, D783–D788 (2020).

51. Dilthey, A. T., Moutsianas, L., Leslie, S. & McVean, G. HLA*IMP—an integrated framework for imputing classical HLA alleles from SNP genotypes. Bioinformatics 27, 968–972 (2011).

52. Patsopoulos, N. A. et al. Fine-mapping the genetic association of the major histocompatibility complex in multiple sclerosis: HLA and non-HLA effects. PLoS Genet. 9, e1003926 (2013).

53. Levin, M. L. The occurrence of lung cancer in man. Acta Unio Int. Contra Cancrum 9, 531–541 (1953).

54. Miettinen, O. S. Proportion of disease caused or prevented by a given exposure, trait or intervention. Am. J. Epidemiol. 99, 325–332 (1974).

55. Choi, S. W. & O’Reilly, P. F. PRSice-2: Polygenic Risk Score software for biobank-scale data. Gigascience 8, (2019).

56. King, T., Butcher, S. & Zalewski, L. Apocrita - High Performance Computing Cluster for Queen Mary University of London. (2017). doi:10.5281/zenodo.438045.

57. Pandit, L. et al. European multiple sclerosis risk variants in the south Asian population. Mult. Scler. 22, 1536–1540 (2016).

58. Jacobs, B. M. et al. Gene-environment interactions in multiple sclerosis: A UK biobank study. Neurology-Neuroimmunology Neuroinflammation 8, (2021).

59. Privé, F. et al. Portability of 245 polygenic scores when derived from the UK Biobank and applied to 9 ancestry groups from the same cohort. Am. J. Hum. Genet. 109, 12–23 (2022).

