## supplementary methods for "Genetic determinants of Multiple Sclerosis susceptibility in diverse ancestral backgrounds"

**Supplementary data for ‘Genetic determinants of Multiple Sclerosis risk in diverse ancestral backgrounds’**

**Overview**

This supplementary file provides additional information regarding the quality control of genetic data used to produce the results in this article. An overview of genetic quality control, including the number of individuals and markers excluded/included at each stage is provided in **supplementary figure 1**. Information regarding the cohort itself can be found in the cohort profile^1^.

**Genotyping & genotype calling**

Saliva samples were obtained from participants using the Oragene OG-600 saliva kits. Genomic DNA was extracted from saliva samples at the Queen Mary University of London Genome Centre according to manufacturer’s instructions. Genomic DNA was genotyped using the Illumina Global Screening Array version 3.0 24-bead chip with additional Multi-Disease content. Genotype calling and quality control were performed using Illumina GenomeStudio version 2.0. Raw intensity files were imported into a single GenomeStudio project. SNP positions were obtained from the publicly-available SNP manifest in genome build hg38.  For initial genotype calling, we used the custom cluster file used for the Genes & Health Study^2^, a genetic cohort study of ~50,000 British people of South Asian ancestry which also genotyped participants using Oragene saliva kits and the Illumina GSA version 3.0 chip with EAMD content. We used an initial no-call threshold of 0.15 (the default).

Prior to filtering there were samples from 829 unique Oragene IDs – note that some individuals participated twice - genotyped at 730,059 markers. We performed initial genotype calling using the G&H cluster file, then performed filtering on genetic markers, excluded copy number variants (CNVs), mitochondrial variants, and Y chromosome variants. We excluded markers with any of the following: low cluster separation (cluster separation score < 0.3), with call frequencies < 97%, with heterozygote mean intensity (R mean) of < 0.2, heterozygote mean theta values of < 0.2 or > 0.8, or minor allele frequency of < 1%. After exclusion of these markers we then recalculated call rates and removed samples with call rate < 97%. We then performed genetic sex inference using the X chromosome, and subsequently removed sex chromosome variants, markers with heterozygote excess of < -0.3 or >0.2, and those with a call frequency of <95%. After a further round of recalculating call rates using only the high-quality retained markers and restricting to samples with call rate >= 97%, where duplicates were available we took the sample with the higher quality genotyping data as determined by the GC50 value – the 50^th^ percentile of genotype quality score for each individual, i.e. the median genotype quality for that person. High-quality genotype calls for 805 unique Oragene IDs were exported into PLINK version 1^3^ binary format, forcing all alleles onto the forward strand. We performed further genotype quality control, restricting to markers which did not deviate from Hardy-Weinberg equilibrium at *P* < 1x10^-10^, yielding a dataset of 259,150 genotyped markers.

#### **Kinship estimation and duplicate removal**

We removed participants without any covariate data (N=46), i.e. participants who returned a saliva kit but did not complete any of the required questionnaire. To identify duplicates, contaminated samples and highly-related samples, we calculated kinship coefficients between individuals in this dataset using KING^4^. We identified 23 duplicates, which reflected participants who took part twice, and retained the higher quality sample for each pair. Of the remaining samples, there were 7 pairs of related above 3^rd^ degree relatedness, which were retained.

#### **Heterozygosity, missingness and sex check**

The mean genotyping rate was >99.9%. We estimated heterozygosity per sample using the complete set of genotyped markers. All individuals fell within a plausible range of *F* statistics (-0.08 to + 0.27) and so none were excluded on this basis. We compared self-stated gender information with genetically-inferred biological sex as a further sense check. Concordance was extremely high – 496 / 497 people (99.8%) identifying as female were of female biological sex, and 190 / 192 self-stated males (99.0%) were of male biological sex. Individuals with a difference between stated gender and biological sex were not excluded.

#### **Genotype imputation**

Imputation was performed using the Haplotype Reference Consortium (HRC) version 1.1 via the Michigan imputation server^5^. Briefly, genotypes for each of the 736 genotyped participants in the post-QC dataset were phased with Beagle (version 5.4)^6^ and imputed with Minimac4^7^ using the reference genomes present in the HRC panel^8^. Following an initial first-pass imputation, we excluded variants which were incompatible with the reference panel and flipped variants which were genotyped on the opposite strand. Imputation quality was high (median R^2^ > 0.75) for common (MAF > 1%) variants (**supplementary figure 2**). We hard-called genotypes based on imputed dosage, and set to missing calls > 0.1 from the nearest integer (i.e. outside of 0±0.l, 1±0.l, or 2±0.l). We performed further QC on the imputed data, restricting to autosomal biallelic SNPs with MAF >= 0.01, imputation R^2^ > 0.3, missingness <10%, and no deviation from HWE at P < 1e-5. The final dataset comprised 736 people with imputed genotypes at 2.7 million SNPs.

#### **Integration with UK Biobank controls**

We combined our case-only cohort with data from UK Biobank to obtain controls and a small number of additional cases (application ID 78867). In brief, UK Biobank is a longitudinal genotype-phenotype cohort study with linked healthcare records, questionnaire data, genotyping, and several other rich phenotyping datasets for ~500,000 British adults recruited between the ages of 40 and 69^9^. As the majority of UKB participants do not have MS this is a rich source of ancestrally-diverse control participants for GWAS. In addition there are also approximately 2,600 people with MS in the cohort (defined as any occurrence of an MS diagnostic code in the electronic healthcare record or self-report; UKB field ID 131043)^10^. We used imputed genetic data (UKB category ID 100319) derived from genotyping on one of two arrays (Axiom or Bileve) followed by imputation to the Haplotype Reference Consortium imputation panel. We performed light first-pass quality control, filtering to markers with low missingness (<10%), no deviation from HWE at P < 1x10^-5^, a MAF of 0.01, and an imputation quality score of >0.3. We excluded individuals with >10% missing genotypes and converted these genotype data to PLINK format (in hg19 co-ordinates).

We identified a common subset of SNPs passing QC and present in the UKB and ADAMS datasets with perfectly compatible SNP positions and alleles (i.e. identical Chromosome:Position:Ref:Alt). We performed further filtering, removing SNPs with a combined MAF of <**1**%, with deviation from HWE at P < 1x10^-10^, or missingness > 10%. To account for possible strand-flips – i.e. SNPs assessed on opposite strands in the two datasets – we removed palindromic SNPs (i.e. A/T or C/G SNPs) – resulting in a dataset of 2.2 million variants.

#### **Ancestry inference**

To infer the genetic ancestry of the cohort, we used the joint callset from two reference datasets containing samples of defined ancestry: the Human Genome Diversity Project (HGDP) and 1,000 Genomes (1KG) Project. Data were downloaded in hg38 format from the GnomAD website (<https://gnomad.broadinstitute.org/>) and converted to hg19. We restricted both the ADAMS-UKB genotypes and HGDP-1KG genotypes to an overlapping SNP set of non-palindromic autosomal markers with compatible alleles and passing quality control in both datasets (MAF >0.01, genotype missingness < 1%, and no deviation from HWE at *P* < 1x^-10^). To ensure the ancestry estimation in the reference cohort was not biased by relatedness, we calculated kinship coefficients between samples in the HGDP-1KG dataset and removed one of each pair of individuals (within the HGDP-1KG dataset) related at a kinship coefficient of 0.0844 or greater (equivalent to 3^rd^ degree relatedness, i.e. a first cousin). We LD-pruned these SNPs with a pruning R^2^ of 0.1. We calculated the top 50 principal components using PLINK and projected the ADAMS-UKB samples onto this same principal component space using the SNP loadings and allele frequencies from the HGDP-1KG samples. Using these principal component scores and the known ancestry labels, we trained a random forest classifier on the HGDP-1KG data to predict genetic ancestry from the first 50 genetic PCs. This classifier had an average accuracy of 99.9% in the training set.

We then applied this random forest classifier to the ADAMS-UKB samples to estimate the ‘best-guess’ continental-level genetic ancestry for each participant in the dataset. Using this approach, we defined three broad ancestral superpopulations (**supplementary table 2**): (Central) South Asian (SAS), African (AFR), and European (EUR; **supplementary figure 3**). We identified individuals with low-confidence ancestry assignments (i.e. maximal probability < 80%), likely reflecting recent admixture (**supplementary figures 4-6**): this threshold was determined heuristically by examining the distribution of inferred probabilities within each self-reported ethnic group in UKB & ADAMS, and by inspecting the distribution of probabilities within the UKB-determined subset of ‘White British’ European-ancestry participants (data field 22006). Inferred ancestry was cross-validated with self-reported ethnicity data, where available (**supplementary figures 7 - 8**). Individuals with low-confidence ancestry assignments were excluded for the purposes of downstream analysis (**supplementary figure 9**).

**Within-ancestry quality control**

We then separated two broad ancestry groups (South Asian [SAS] and African [AFR]) and performed within-ancestry quality control by excluding variants which were rare (MAF <1%), deviated from HWE (P<1x10^-20^), or were missing at >10%. To identify variants which might confer bias due to differences in the generation of genetic data from UKB and ADAMS (e.g. DNA extraction, genotyping array, imputation), we performed a case vs case GWAS of MS cases in ADAMS vs UKB within each ancestral group, taking advantage of the presence of participants with MS in both datasets. We excluded SNPs which differed by > 0.2 in allele frequency, had a MAF of <1% in both cohorts, an allele count of 0 in both cohorts, or which showed evidence of batch effect (Fisher’s exact test *P* < 0.05 for association with cohort of origin; **supplementary figure 10**). We performed another round of PCA on a pruned set of non-MHC markers (pruning R^2^ 0.05, MAF >= 10%, missingness < 1%) within each ancestral cluster and excluded samples lying outside of 3 standard deviations from the mean of any of the first ten principal components. To boost the number of testable genetic markers, we then jointly reimputed both the SAS-ancestry and AFR-ancestry cohorts using the 1,000 genomes reference data (multi-ancestry, N = 2,504). We jointly phased haplotypes using EAGLE (v2.4.1)^11,12^ and imputed using Minimac4. Following re-imputation, we hard-called genotypes using a threshold of 0.49 (i.e. imputed dosages 0 – 0.49 were called as 0, 0.51 – 1.49 were called as 1, and 1.51 – 2 were called as 2), and performed further quality control, restricting to autosomal, biallelic, common (MAF > 5%) well-imputed SNPS (R^2^ > 0.7) with low missingness (<10%) and no strong deviation from HWE (P > 1x10^-50^). Finally, we excluded individuals with >10% missing genotypes within the re-imputed data, and we performed a further round of excluding individuals lying >3 standard deviations from the mean of any of the first ten PCs. Following this procedure, there was no visual separation of cases and controls in the first ten PCs (**supplementary figures 10 & 11**).

**References**

1.	Jacobs, B. M. *et al.* Cohort profile: ADAMS project: a genetic Association study in individuals from Diverse Ancestral backgrounds with Multiple Sclerosis based in the UK. *BMJ Open* **13**, (2023).

2.	Finer, S. *et al.* Cohort Profile: East London Genes & Health (ELGH), a community-based population genomics and health study in British Bangladeshi and British Pakistani people. *Int. J. Epidemiol.* **49**, 20–21i (2020).

3.	Chang, C. C. *et al.* Second-generation PLINK: rising to the challenge of larger and richer datasets. *Gigascience* **4**, 7 (2015).

4.	Manichaikul, A. *et al.* Robust relationship inference in genome-wide association studies. *Bioinformatics* **26**, 2867–2873 (2010).

5.	Das, S. *et al.* Next-generation genotype imputation service and methods. *Nat. Genet.* **48**, 1284–1287 (2016).

6.	Browning, B. L., Tian, X., Zhou, Y. & Browning, S. R. Fast two-stage phasing of large-scale sequence data. *Am. J. Hum. Genet.* **108**, 1880–1890 (2021).

7.	Fuchsberger, C., Abecasis, G. R. & Hinds, D. A. Minimac2: Faster genotype imputation. *Bioinformatics* **31**, 782–784 (2015).

8.	McCarthy, S. *et al.* A reference panel of 64,976 haplotypes for genotype imputation. *Nat. Genet.* **48**, 1279–1283 (2016).

9.	Bycroft, C. *et al.* The UK Biobank resource with deep phenotyping and genomic data. *Nature* **562**, 203–209 (2018).

10.	Jacobs, B. M. *et al.* Gene-Environment Interactions in Multiple Sclerosis: A UK Biobank Study. *Neurol Neuroimmunol Neuroinflamm* **8**, (2021).

11.	Loh, P.-R., Palamara, P. F. & Price, A. L. Fast and accurate long-range phasing in a UK Biobank cohort. *Nat. Genet.* **48**, 811–816 (2016).

12.	Loh, P.-R. *et al.* Reference-based phasing using the Haplotype Reference Consortium panel. *Nat. Genet.* **48**, 1443–1448 (2016).
