## supplementary figures for "Genetic determinants of Multiple Sclerosis susceptibility in diverse ancestral backgrounds"

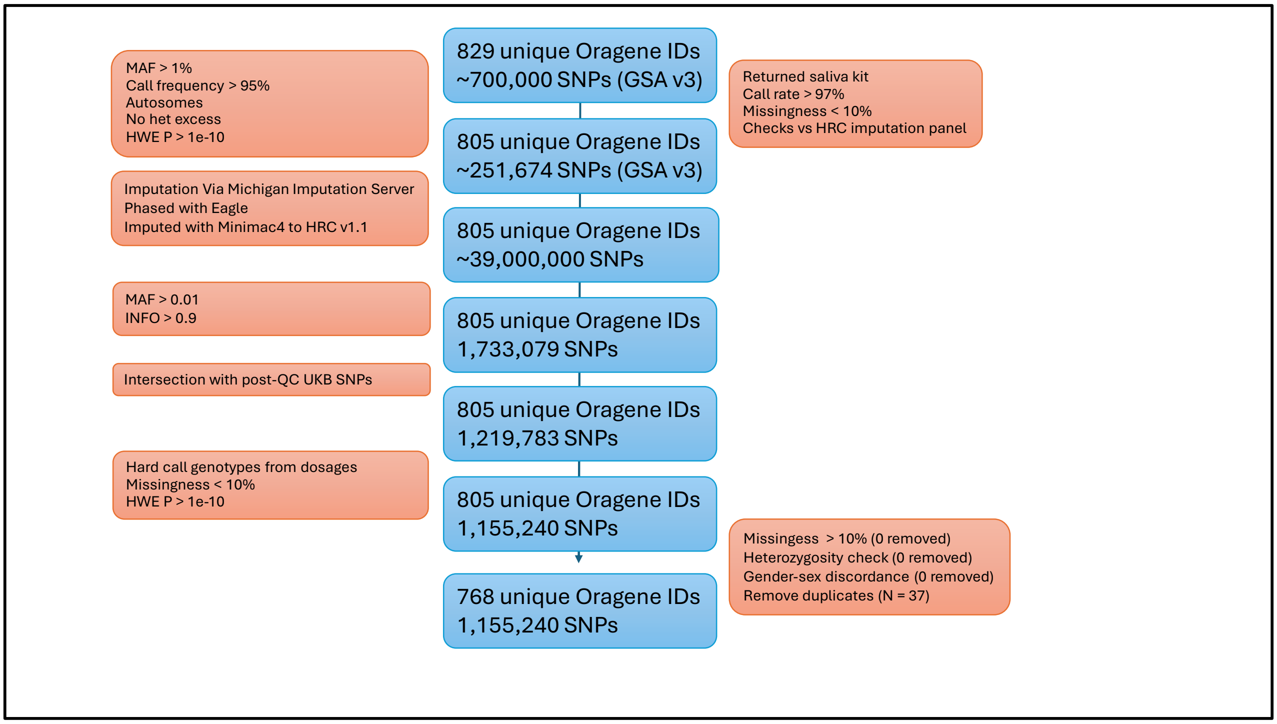


Supplementary figure 1: flowchart showing SNP & individual QC of initial data.

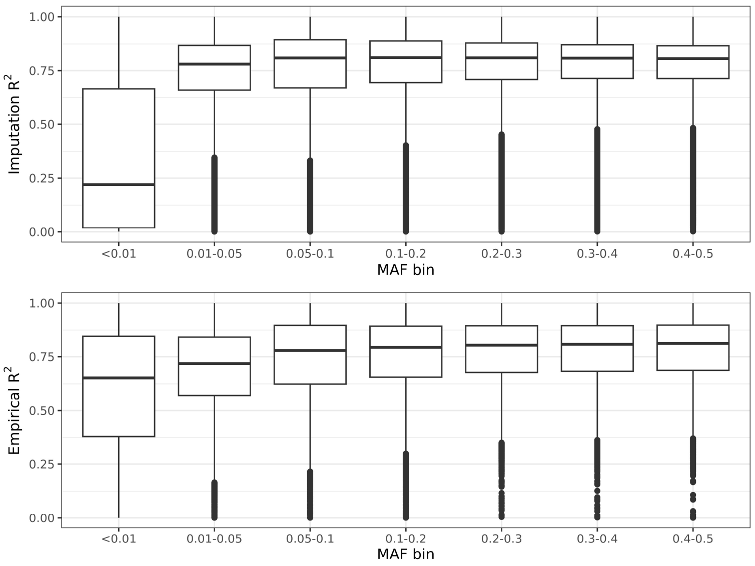


Supplementary figure 2: Imputation quality vs MAF in the ADAMS cohort

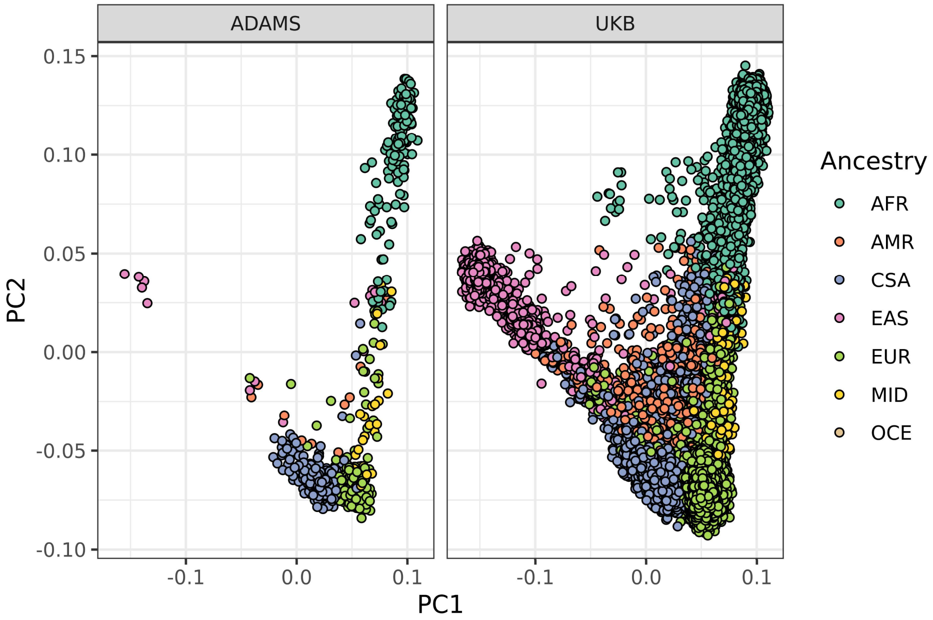


Supplementary figure 3: PCA plot of ADAMS & UKB following ancestry inference. Ancestry key: AFR = African, AMR = Amerindian, CSA (referred to as SAS elsewhere) = Central South Asian / South Asian, EAS = East Asian, EUR = European, MID = Middle Eastern, OCE = Oceanian.

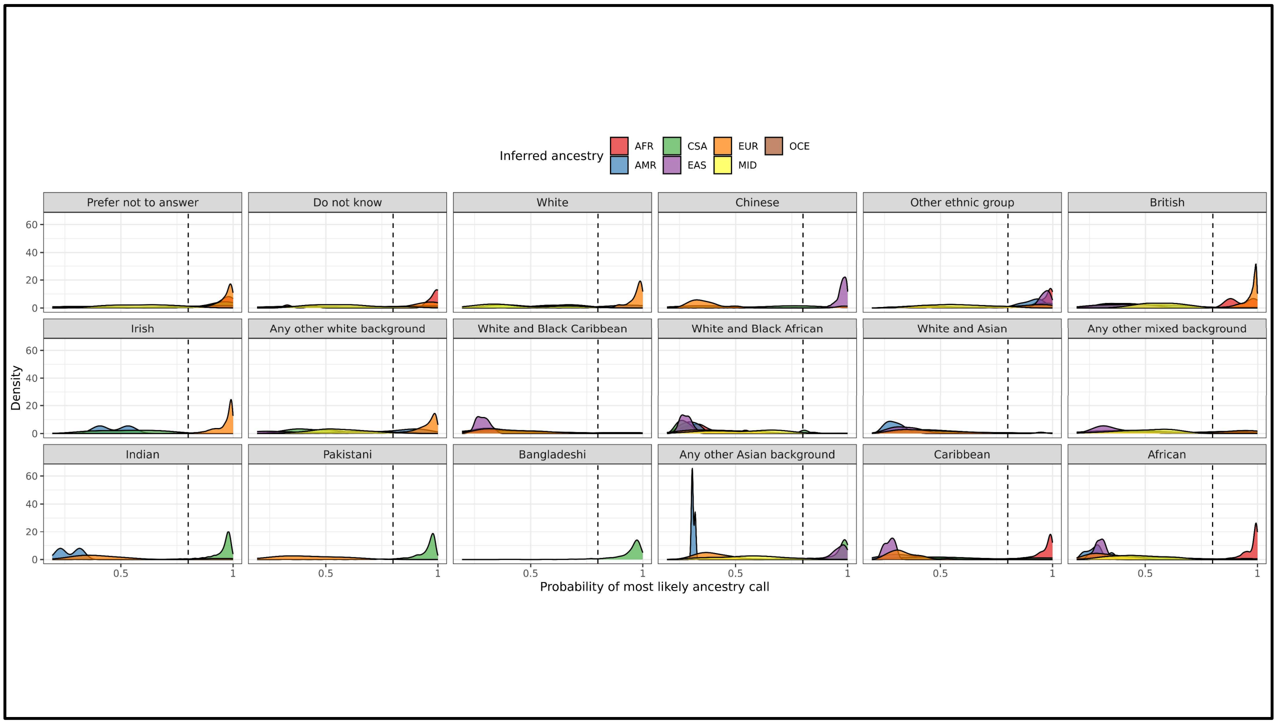


Supplementary figure 4: Probability of genetically-inferred ancestry calls vs self-reported ethnicity in UK Biobank. The dotted line shows the arbitrary cutoff of 80%. Ethnicities with N > 200 are shown. Ancestry key: AFR = African, AMR = Amerindian, CSA (referred to as SAS elsewhere) = Central South Asian / South Asian, EAS = East Asian, EUR = European, MID = Middle Eastern, OCE = Oceanian.

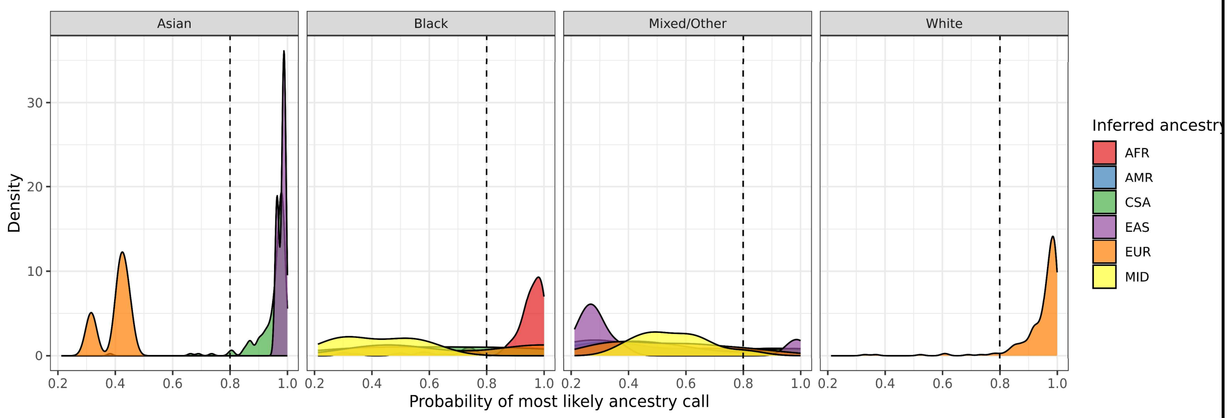


Supplementary figure 5: Probability of genetically-inferred ancestry calls vs self-reported ethnicity in ADAMS. The dotted line shows the arbitrary cutoff of 80%. Ancestry key: AFR = African, AMR = Amerindian, CSA (referred to as SAS elsewhere) = Central South Asian / South Asian, EAS = East Asian, EUR = European, MID = Middle Eastern, OCE = Oceanian.

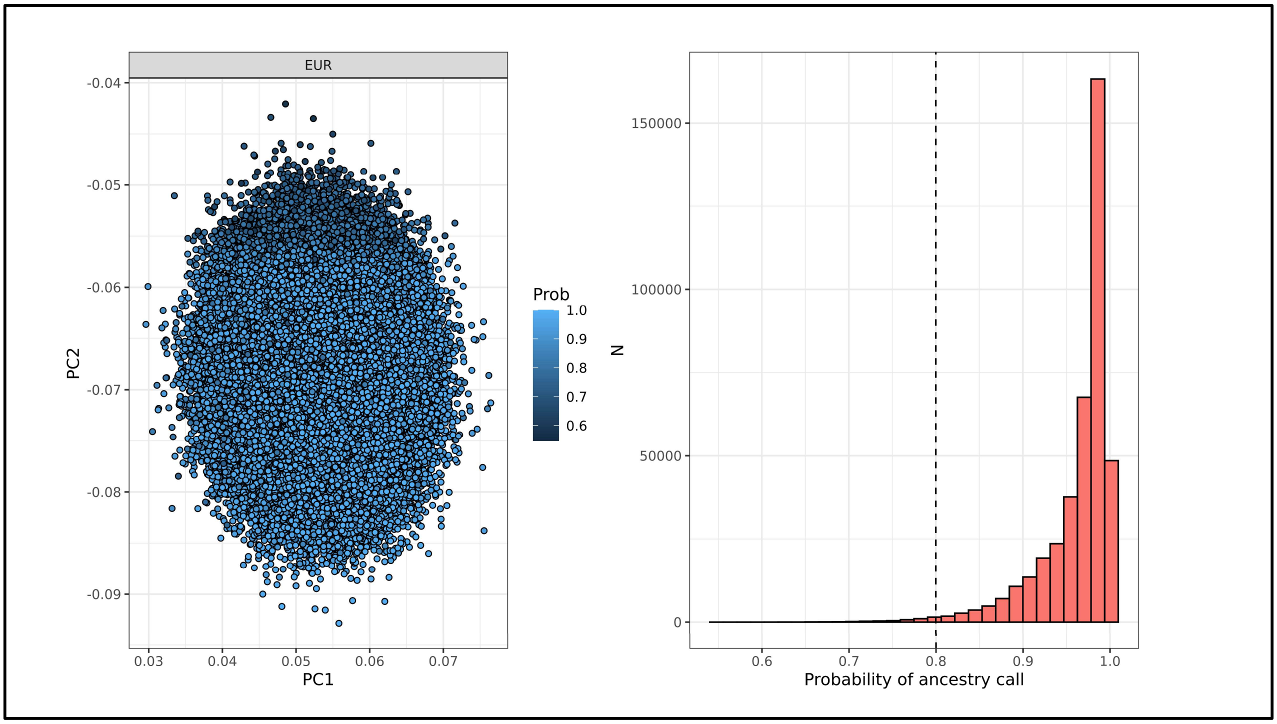


Supplementary figure 6: PCA plot and histogram showing the genetically inferred ancestry & probability of UKB-identified genetic Caucasians (UKB field genetic ethnic grouping)

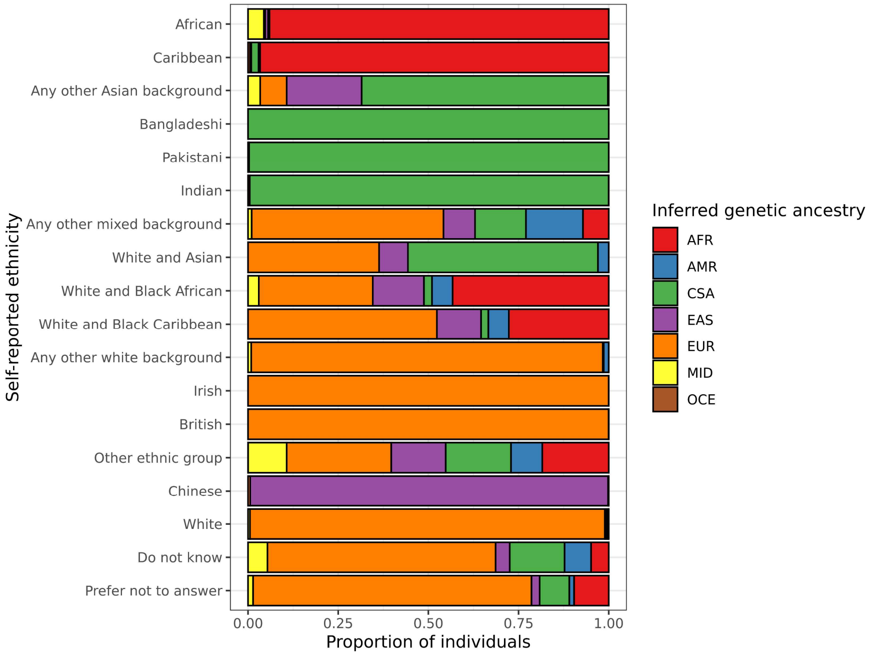


Supplementary figure 7: Concordance between self-reported ethnicity and inferred genetic ancestry in UKB. Ancestry key: AFR = African, AMR = Amerindian, CSA (referred to as SAS elsewhere) = Central South Asian / South Asian, EAS = East Asian, EUR = European, MID = Middle Eastern, OCE = Oceanian.

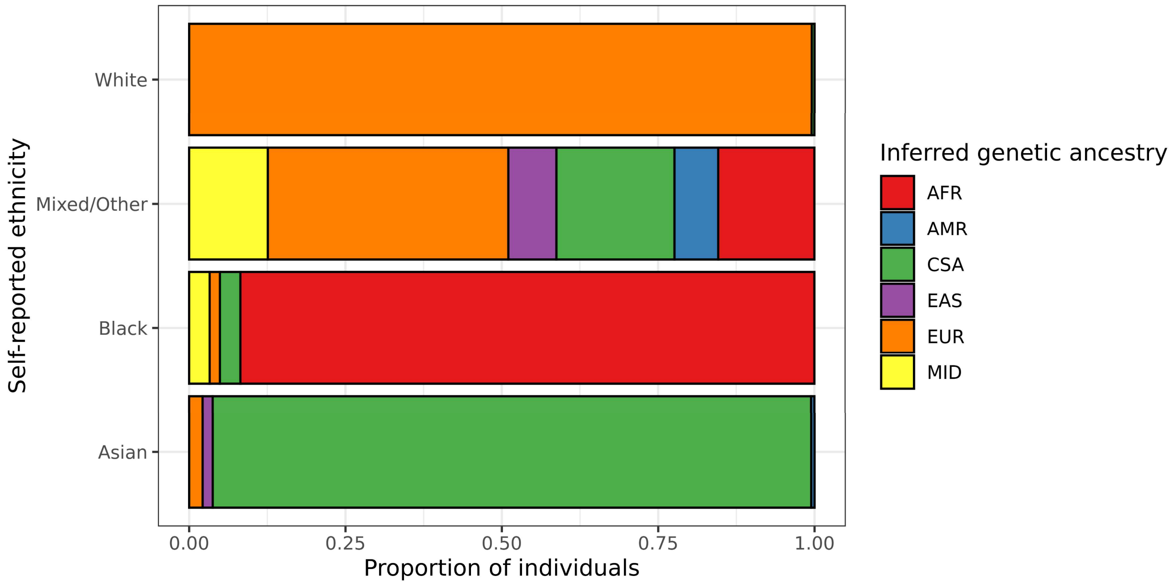


Supplementary figure 8: Concordance between self-reported ethnicity and inferred genetic ancestry in ADAMS. Ancestry key: AFR = African, AMR = Amerindian, CSA (referred to as SAS elsewhere) = Central South Asian / South Asian, EAS = East Asian, EUR = European, MID = Middle Eastern, OCE = Oceanian.

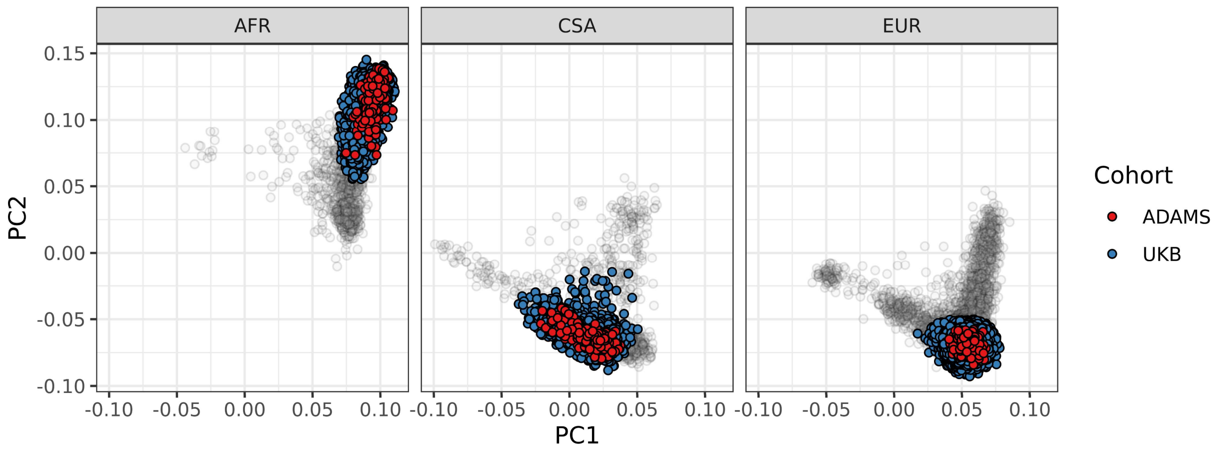


Supplementary figure 9: PCA plots showing ADAMS-UKB combined samples for the 3 major ancestry groups, with ambiguous calls (<80% probability) greyed out. Ancestry key: AFR = African, CSA (referred to as SAS elsewhere) = Central South Asian / South Asian, EUR = European.

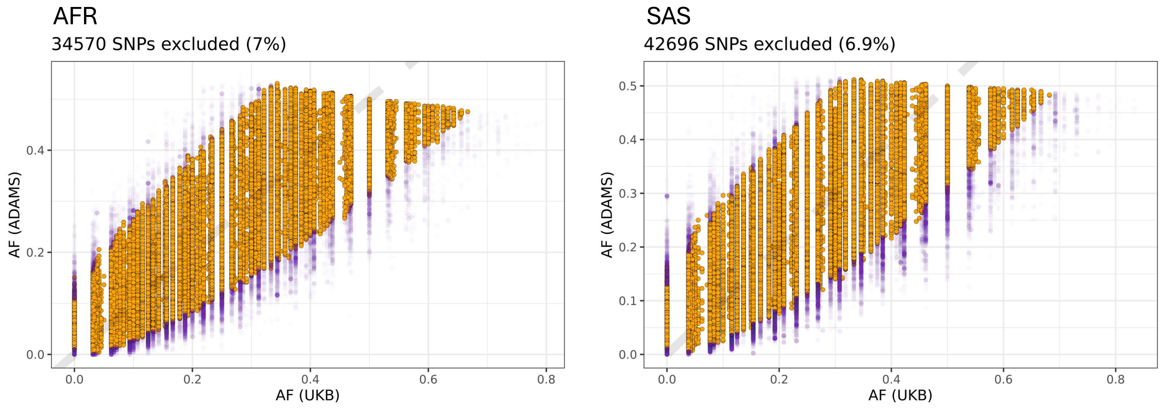


Supplementary figure 10: Frequency plots showing the frequency of variants in ADAMS cases vs UKB cases of similar inferred ancestry. Each point is a variant. Purple points denote excluded variants. Variants were excluded if: their frequency differed by >0.2 between the cohorts, the Fisher's exact test P value for association with cohort was < 0.05, the allele was not observed in either cohort, or the allele was present at a frequency of <1% in both cohorts. AFR = African ancestry, SAS = South Asian ancestry.

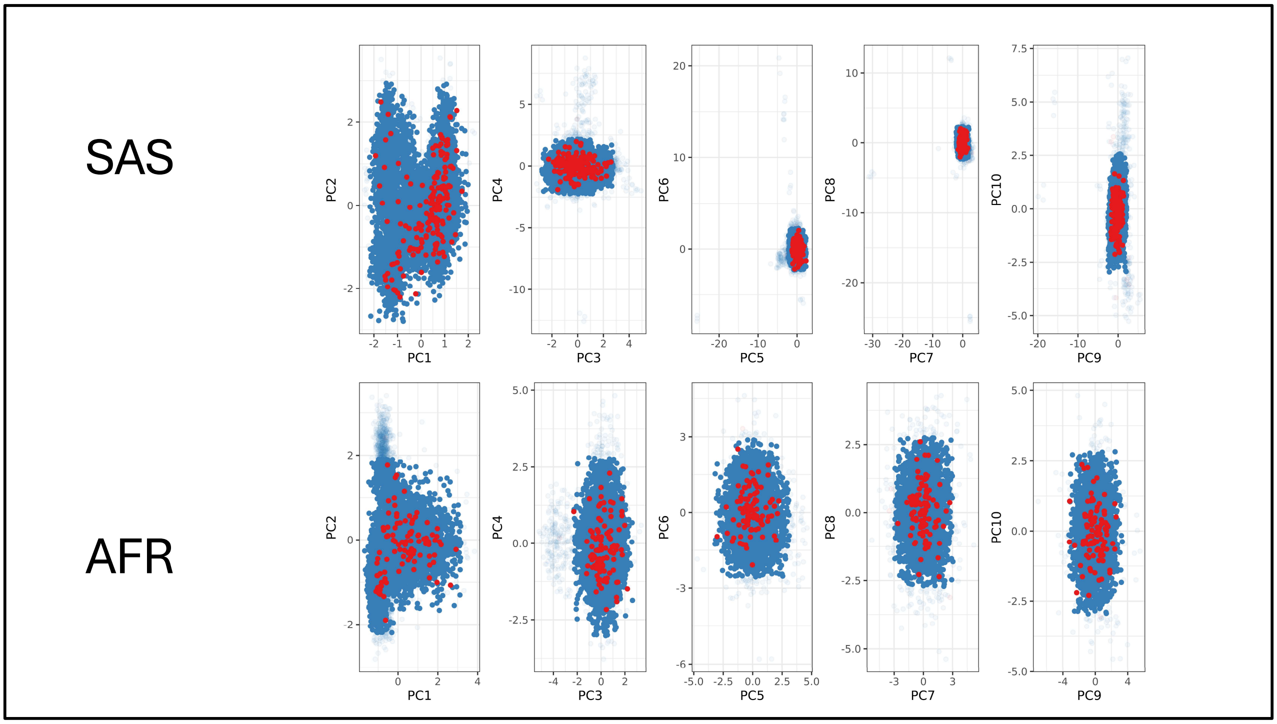


Supplementary figure 11: PC plots (PCs 1 – 10) following joint reimputation showing cases in red and controls in blue. PCA outliers (excluded points) are shown as translucent points. There was no clear separation of cases & controls. AFR = African ancestry, SAS = South Asian ancestry.

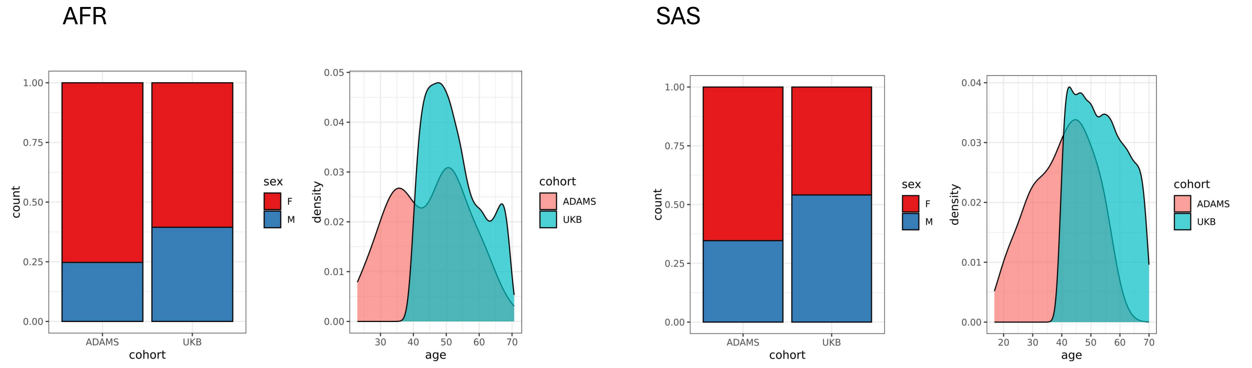


Supplementary figure 12: barplots showing the self-stated gender distributions in UKB vs ADAMS. Density plots show the distribution of age at recruitment. The African-ancestry cohort is shown on the left, and the South Asian-ancestry cohort on the right. The ADAMS cohort had a higher proportion of women and was substantially younger than UKB. AFR = African ancestry, SAS = South Asian ancestry.

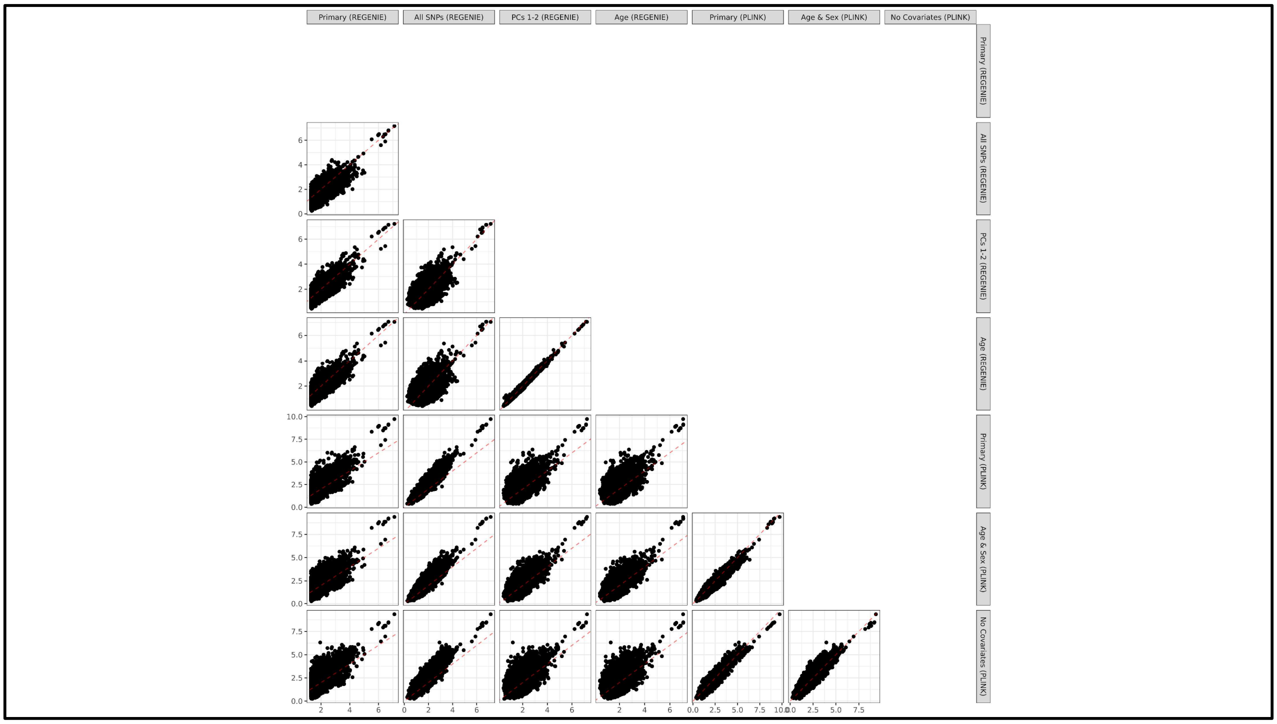


Supplementary figure 13: comparison of –log10(P) values from various GWAS models & sensitivity analyses in AFR-ancestry.

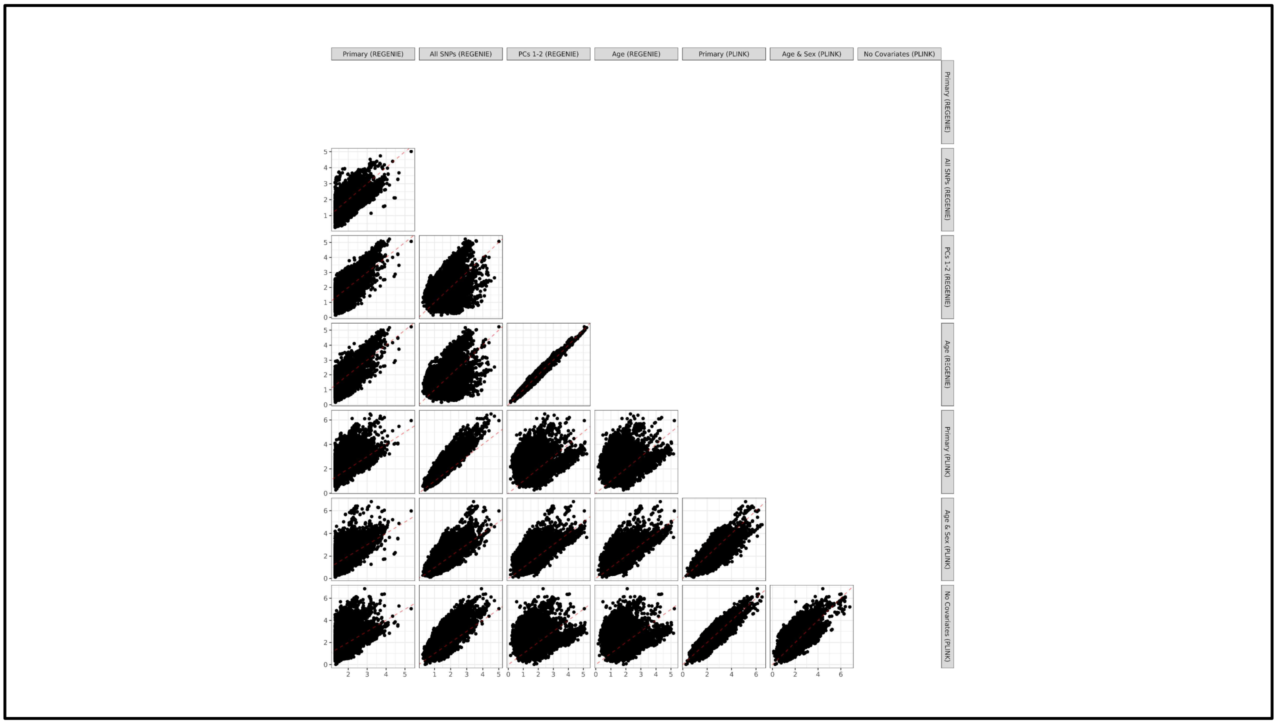


Supplementary figure 14: comparison of –log10(P) values from various GWAS models & sensitivity analyses in SAS-ancestry.

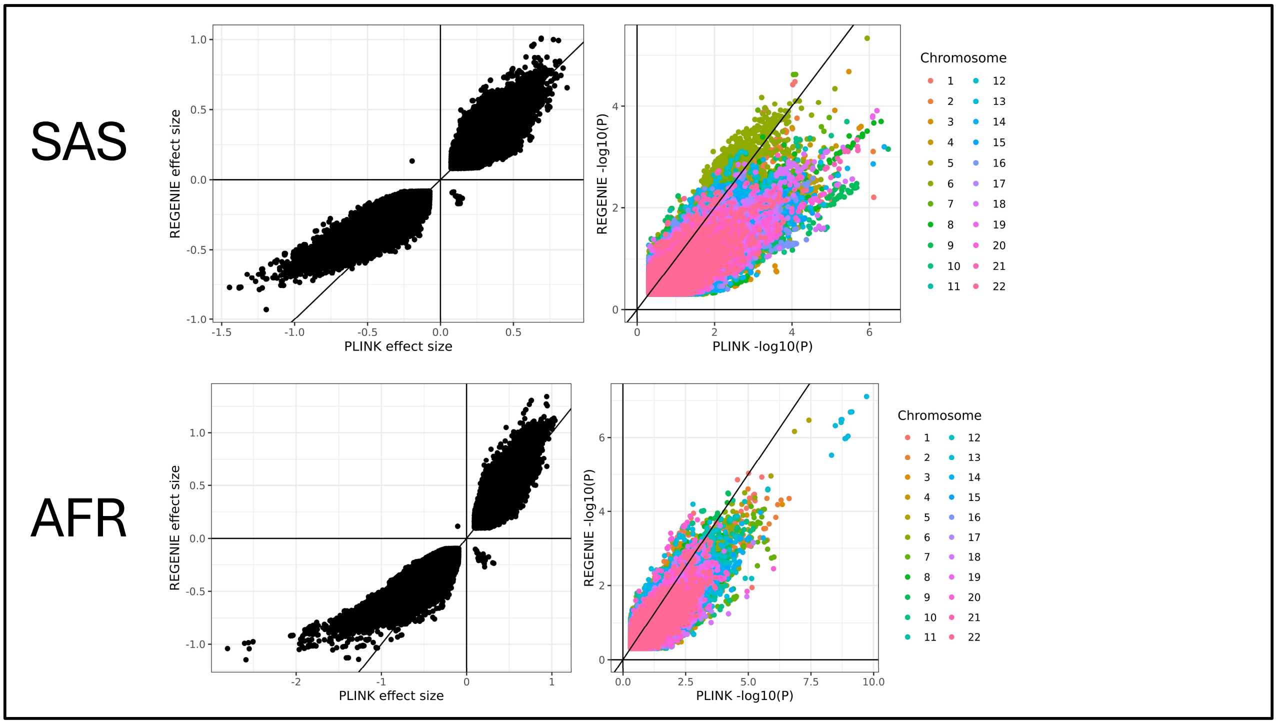


Supplementary figure 15: comparison of effect sizes (left panels) and –log10(P) values (right panels) from fixed effects GWAS (PLINK, x axis) vs random effects GWAS (REGENIE, y axis), showing more conservative P values and effect sizes with REGENIE. The top and lower panels show the results for the SAS (top) and AFR (bottom) cohorts. AFR = African ancestry, SAS = South Asian ancestry.

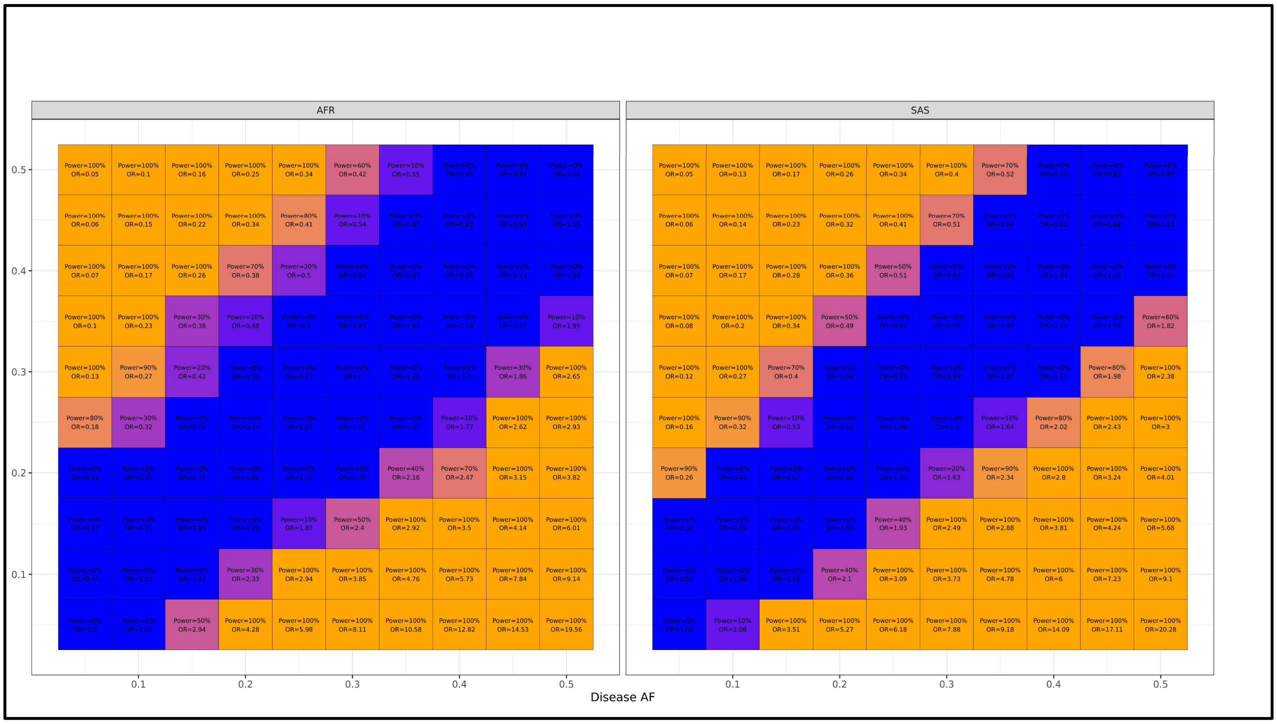


Supplementary figure 16: power calculations indicating the power to detect a genome-wide significant signal given the number of cases & controls in each ancestry across a range of allele frequency differences. AFR = African ancestry, SAS = South Asian ancestry.
